# Pulmonary extracellular vesicles drive alveolar macrophage dysfunction via microRNA transfer in Acute Respiratory Distress Syndrome

**DOI:** 10.64898/2026.06.13.26355564

**Authors:** Katie L. Spencer, Charlie Mafham, Joshua Price, Ellen Jenkins, Celine H. Chen, Samuel Quarton, Louise E. Crowley, Xiaohong Jiang, Jose R. Hombrebueno, Michael A. Matthay, Mark Lindsay, Babu Naidu, David R. Thickett, Dhruv Parekh, Aaron Scott, Rahul Y. Mahida

## Abstract

**Background:** Alveolar macrophage (AM) dysfunction contributes to Acute Respiratory Distress Syndrome (ARDS) pathogenesis. We investigated the role of extracellular vesicles (EVs) in mediating this dysfunction.

**Methods:** Pulmonary EVs were isolated from broncho-alveolar lavage and non-directed bronchial lavage samples of ventilated sepsis patients with and without ARDS, and post-operative control patients via ultracentrifugation. AMs were isolated from lung tissue resections of lobectomy patients. AMs were treated with pooled EVs for 24 hours prior to functional, metabolic and autophagy profiling. EV cargo was profiled via small RNA transcriptomics and proteomics. Mechanistic role of EV microRNAs was assessed via mimic / antagomir transfection.

**Results:** Pulmonary EVs from sepsis patients with ARDS impaired AM efferocytosis, and control EVs had no effect. ARDS EV treatment enhanced AM mitochondrial-linked respiration, but not glycolysis. ARDS EV treatment impaired LC3B-II and LAMP1 expression, indicating dysregulated AM autophagy-lysosomal machinery. Proteomics revealed downregulation of innate immune pathways in ARDS EVs. Transcriptomics revealed enrichment of 24 microRNAs in ARDS EVs; miR-652-3p was the most enriched, validated by RT-qPCR. EV miR-652-3p was associated with 90-day mortality (9.20 vs 0.59 RQ, p=0.0295) and inversely correlated with oxygenation (PaO_2_/FiO_2_). AM transfection with miR-652-3p mimic induced similar dysregulation of function and autophagy as ARDS EVs. Transfection of ARDS EVs with antagomirs to miR-652-3p prior to AM treatment partially rescued efferocytosis and autophagy.

**Conclusions:** Targeting EV miR-652-3p may restore alveolar macrophage function and reduce excessive inflammation, thus offering a novel therapeutic strategy for patients with ARDS.

## Introduction

Acute Respiratory Distress Syndrome (ARDS) is an inflammatory pulmonary disorder, commonly precipitated by sepsis. Neutrophilic inflammation and alveolar-capillary barrier damage cause alveolar oedema and refractory hypoxia, requiring prolonged mechanical ventilation. The mainstay of treatment remains supportive, and mortality is persistently high at 40% (1, 2). Thus, there is an urgent need to identify novel therapeutic targets for ARDS.

Alveolar macrophages (AMs) co-ordinate pulmonary immune responses and undertake key pro-resolving functions including efferocytosis (clearance of apoptotic cells). ARDS is associated with massive neutrophil influx into alveoli. As these neutrophils become apoptotic, they require efficient clearance to prevent secondary necrosis. This process releases inflammatory mediators within the alveolar space, contributing to tissue damage and exaggerated inflammation in ARDS (3). We have previously reported that patients with sepsis-related ARDS have impaired AM efferocytosis and increased pulmonary neutrophil apoptosis compared to ventilated sepsis patients without ARDS (4). Impaired AM efferocytosis is associated with increased inflammatory cytokine release, 30-day mortality and duration of mechanical ventilation (4). We have also shown that AMs from ARDS patients have an impaired ability to metabolise cortisone, indicating a global defect in AM function and associated metabolic dysregulation (5). Treatment of healthy AMs with broncho-alveolar lavage (BAL) fluid from ARDS patients impairs efferocytosis, reproducing the same functional defect observed in ARDS (6). Thus, components of ARDS BAL must be responsible for the dysfunction observed.

Extracellular vesicles (EVs) are membrane-bound anuclear structures which constitute an intercellular communication mechanism, allowing transfer of biologic cargo (mitochondria, RNA) between cell types (7). EV uptake can alter gene expression within cells. Pathogenic EVs are released when *ex vivo* perfused human lungs are injured with E.coli; these EVs then mediate inflammatory injury when isolated and administered to uninjured perfused human lungs (8). Murine models of ARDS show that EV transfer of microRNA (miR) cargo to alveolar macrophages (AMs) can increase inflammatory cytokine release (9). ARDS models have also shown that EVs are predominantly taken up within the alveoli by tissue-resident AMs via MerTK (10). EV transfer of miRNA to murine macrophages alters metabolic profile and function (11). We have previously reported that CD14^+^/CD81^+^ BAL EVs are enriched in patients with sepsis-related ARDS; this EV subset is also associated with increased 30-day mortality (12). Thus, EVs may contribute to ARDS pathogenesis by promoting inflammation.

In this study, we investigated whether the pulmonary EVs are responsible for dysregulating AM function and metabolism in sepsis-related ARDS. We also mechanistically evaluated the role of EV microRNA cargo on mediating these effects.

## Results

### Characterisation of pulmonary EVs in sepsis patients with ARDS

The demographic and clinical characteristics of ARDS and control patients are described in Table 1. In the AM-ARDS study 14 patients with ARDS and 14 patients without ARDS (sepsis control) were recruited; all had BAL collected. In the VESPER study 15 patients with ARDS and 12 patients without ARDS (sepsis control) were recruited; all had non-directed bronchial lavage (NDBL) collected. BAL samples taken post-operatively from 13 patients enrolled to the Vindaloo trial were used as an additional ventilated control group (13). EVs were isolated from the BAL and NDBL samples of all patients and characterised via multiple modalities. Transmission electron microscopy confirmed the presence and morphology of pulmonary EVs (Figure 1A). Nanoparticle tracking analysis (NTA) of BAL EV samples showed that sepsis patients with ARDS had a greater concentration of EVs compared to post-operative controls (Figure 1B, medians 3.13×10^9^ vs 1.65×10^10^/ml, p=0.0088). EV size profiling via NTA confirmed our previous findings that the majority of BAL and NDBL EVs across all patient groups are smaller than 200nm in diameter (Figure 1C-D). Using single particle interferometric reflectance image sensing (SPIRIS - Exoview), we have previously shown that BAL samples from all 3 patient groups contain EVs derived from monocytes, neutrophils, platelets, epithelial and endothelial cells (12). This prior work also revealed enrichment of CD14^+^/CD81^+^ monocyte-derived BAL EVs in sepsis patients with ARDS compared to sepsis patients without ARDS, and an association between this EV subset and increased 30-day mortality in ARDS patients (12). Exoview analysis of NDBL samples showed no significant difference in CD66b^+^ (neutrophil-derived), EPCAM^+^ (epithelial-derived) and CD14^+^ (monocyte-derived) EV populations between sepsis patients with and without ARDS (Supplemental Figure 1A-C). A trend towards elevated CD81^+^/CD14^+^ NDBL EVs in ARDS patients was observed, however this did not reach significance (Supplemental Figure 1C). Total CD81+ EVs were elevated in the NDBL EVs of sepsis patients with ARDS compared to sepsis patients without ARDS (Figure 1E, medians 10,093 vs 16,833, p=0.0241). After isolating BAL and NDBL EVs via ultracentrifugation, and pooling across patient groups, NTA was undertaken to measure pooled EV concentration prior to treatment of AMs (Figure 1F). NTA also confirmed ultracentrifugation as an effective method of isolating EVs, as there were no EVs detected in the BAL supernatant following ultracentrifugation. Our use of 3 complementary techniques to characterise EVs in this study (TEM, NTA and Exoview) aligns with current recommendations to utilise at least 2 EV characterisation methods, as outlined in international guidelines (14).

**Figure 1:**
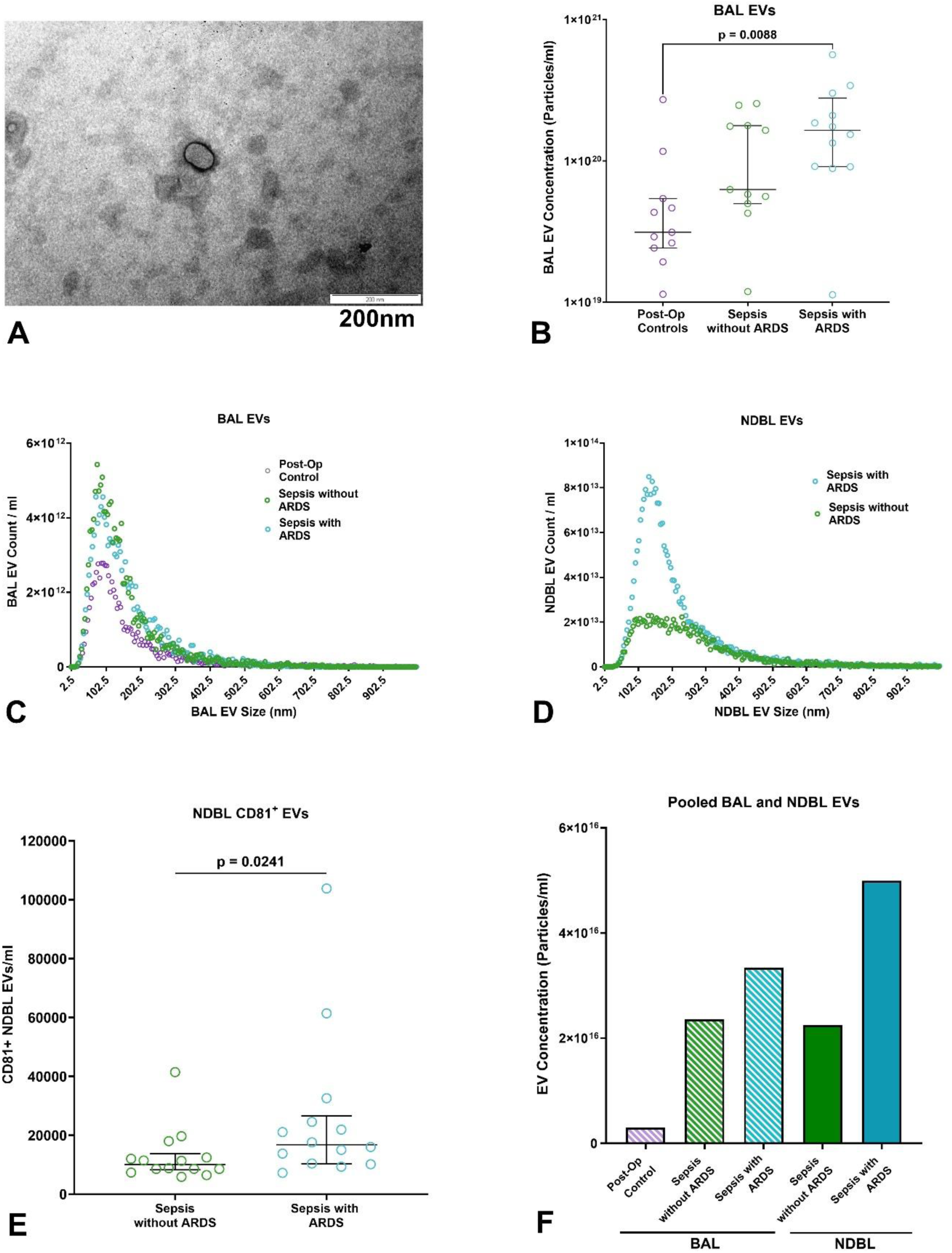
Pulmonary EV characterisation. **A**: Representative transmission electron microscopy image of a BAL EV from a patient with sepsis-related ARDS. **B**: Concentration of BAL EVs as measured by NTA across all 3 patient groups, n≥13 per group. **C**: Size distribution of BAL EV as measured by NTA across all 3 patient groups, medians shown, n≥13 per group. **D**: Size distribution of NDBL EV as measured by NTA across all 3 patient groups, medians shown, n≥5 per group **E**: CD81^+^ NDBL EVs in sepsis patients with and without ARDS, measured by Exoview, n≥12 per group. **F**: Concentration of EVs following isolation from BAL / NDBL and pooling across patient groups, as measured by NTA. BAL: broncho-alveolar lavage, EV: extracellular vesicle, NDBL: non-directed bronchial lavage, NTA: nanoparticle tracking analysis.

**Table 1:** Patient demographics and clinical characteristics.

|  | **BAL Sampling** |  |  | **NDBL Sampling** |  |
| --- | --- | --- | --- | --- | --- |
|  | **AM-ARDS Sepsis with ARDS (n=14)** | **AM-ARDS Sepsis without ARDS (n=14)** | **Post-operative controls (n=13)** | **VESPER Sepsis with ARDS (n=15)** | **VESPER Sepsis without ARDS (n=12)** |
| **Age** | 60.7 (14.1) | 55.1 (16.3) | 66.9 (7.5) | 59.7 (10.3) | 56.9 (15.3) |
| **Male Sex, n (%)** | 10 (71%) | 11 (65%) | 10 (71%) | 8 (53%) | 9 (75%) |
| **Ethnicity: White, n (%)** | 14 (100%) | 14 (100%) | 11 (79%) | 15 (100%) | 11 (92%) |
| **Pneumonia as source of sepsis, n (%)** | 13 (93%) | 10 (71%) | N/A | 14 (93%) | 11 (92%) |
| **P/F ratio on admission** | 22.6 (4.9) | 23.1 (6.7) | N/A | 21.0 (6.37) | 27 (13.6) |
| **APACHE-II score** | 17.1 (7.1) | 14.6 (7.8) | N/A | 20.1 (5.2) | 21.9 (6.9) |
| **SOFA score on admission\*** | 11.5 (8.5 – 15.0) | 10.5 (8.8 - 11.3) | N/A | 12.0 (11.0 – 14.0) | 12.0 (9.5 – 13.8) |
| **ICU LoS in days\*** | 27.0 (15.0-36.0) | 12.0 (7.5 – 19.0) | N/A | 19.0 (9.0 – 28.0) | 14.5 (9.3 – 25.0) |
| **30-day mortality, n (%)** | 3 (21.4%) | 3 (21.4%) | N/A | 5 (33.3%) | 4 (33.3%) |
| **Ventilator-free days to day 28\*** | 8.4 (9.4) | 16.4 (8.7) | N/A | 8.9 (9.8) | 10.2 (9.6) |
| **BAL / NDBL leukocyte count\* x10<sup>6</sup>** | 15.4 (7.3 – 31.3) | 6.4 (3.8 – 27.0) | 8.0 (2.4 – 25.1) | 14.9 (4.0 – 65.2) | 12.3 (4.8 – 17.5) |
| **BAL / NDBL % neutrophils** | 64.9 (28.9) | 49.0 (30.6) | 7.4 (12.0) | 76.2 (18.5) | 74.6 (22.6) |

Values are means (SD) unless otherwise indicated. \*Median and IQR. BAL = broncho-alveolar lavage, ICU = Intensive Care Unit, LoS = length of stay, NDBL = non-directed bronchial lavage, P/F ratio = PaO<sub>2</sub>/FiO<sub>2</sub> ratio, SOFA = sequential organ failure assessment

### Impact of ARDS patient EVs on AM function

AM function was assessed following treatment with pooled EVs. Pooled BAL and NDBL EVs from all patient groups were internalised by AMs (Figure 2A, Supplemental figure 2A). Pooled EV uptake had no impact on AM apoptosis or viability (Supplemental Figure 2B-C). BAL EVs from sepsis patients with ARDS impaired AM efferocytosis compared to EV-depleted BAL (Figure 2C, median fold change 0.546 vs 0.873 p=0.0373). However, BAL EVs from sepsis patients with ARDS had no impact on AM bacterial phagocytosis (Supplemental Figure 2D). NDBL EVs from sepsis patients with ARDS also impaired AM efferocytosis compared to EV-depleted NDBL (Figure 2D, median fold change 0.436 vs 0.738, p=0.0485). BAL EVs from sepsis patients without ARDS and post-operative control patients had no impact on AM efferocytosis (Figure 2E-F). NDBL EVs from sepsis patients without ARDS also had no impact on AM efferocytosis (Figure 2G). Electroporation of ARDS patient EVs with RNAse prior to AM treatment partially rescued efferocytosis (Figure 2F), however electroporation with Proteinase K had no effect (Figure 2G). In summary, the EV component of ARDS patient BAL / NDBL fluid drives the impairment in AM efferocytosis. EV RNA cargo may play a greater role in mediating this effect on AMs compared to protein cargo.

**Figure 2:**
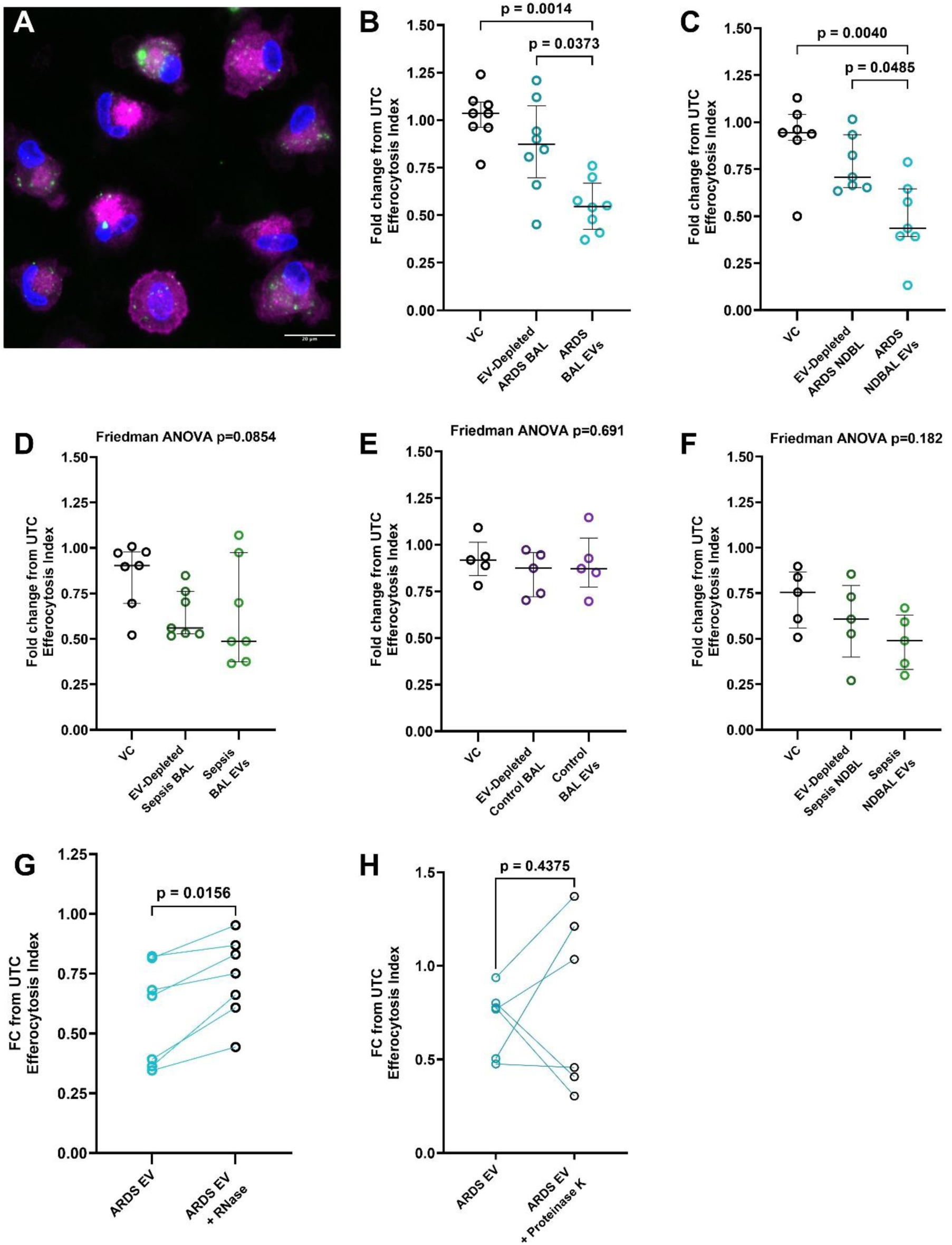
Pulmonary EVs on alveolar macrophage function. Lobectomy AMs were treated with pooled EVs for 24 hours prior to functional assessment. **A**: Representative image of ARDS EV uptake by alveolar macrophages (100x, blue – DAPI, green – celltracker green labelled EVs, purple – CD63). **B**: Impact of pooled BAL EVs and EV-depleted BAL from sepsis patients with ARDS on AM efferocytosis, (n=8 AMs from lobectomy patients). **C:** Impact of pooled NDBL EVs and EV-depleted NDBL from sepsis patients with ARDS on AM efferocytosis, n=7. **D:** Impact of pooled BAL EVs and EV-depleted BAL from sepsis patients without ARDS on AM efferocytosis, n=7. **E:** Impact of pooled BAL EVs and EV-depleted BAL from post-operative control patients on AM efferocytosis, n=5. **F:** Impact of pooled NDBL EVs and EV-depleted NDBL from sepsis patients without ARDS on AM efferocytosis, n=5. **G:** Impact of electroporating pooled ARDS patient EVs with RNase prior to treatment on AM efferocytosis, n=7. **H:** Impact of electroporating pooled ARDS patient EVs with proteinase K prior to treatment on AM efferocytosis, n=7. AM: Alveolar macrophage, BAL: broncho-alveolar lavage, EV: extracellular vesicle, NDBL: non-directed bronchial lavage, UTC: Untreated control, VC: vehicle control (50% saline).

### Impact of ARDS patient EVs on AM metabolic profile

Baseline AM metabolic profiling reveals reliance on mitochondrial respiration (measured by oxygen consumption rate [OCR]) to generate ATP and minimal glycolytic activity (measured by proton efflux rate [PER]) (Supplemental Figure 3A). As previously shown (15), stimulation of AMs with inflammatory mediators (LPS and IFNγ) upregulates OCR, but not PER (Supplemental Figure 3B-C). This metabolic response is divergent to that of monocyte-derived macrophages, which upregulate glycolysis during inflammation (16). Pooled BAL EVs from ARDS patients increased basal (Figure 3A, p=0.0156) and maximal OCR of AMs (Figure 3B, p=0.0273), but had no effect on PER (Figure 3C). These treated AMs demonstrated increased total ATP production (Figure 3D, p=0.0078), which was attributed to increased respiration (Figure 3E, p=0.0078), not glycolysis (Figure 3F). The same effects on AM OCR and PER were observed following treatment with pooled NDBL EVs from ARDS patients (Supplemental Figure 3E-F). Pooled BAL EVs from sepsis patients without ARDS and from post-operative controls had no significant impact on AM OCR (Supplemental Figure 3G). The increase in AM mitochondrial bioenergetics following treatment with ARDS patient BAL-derived EVs occurred independently of changes in mitochondrial mass as assessed by TOMM20 expression (Figure 3F) and membrane potential as evaluated by JC1 assay (Figure 3H). Therefore, the increased AM mitochondrial bioenergetics were unlikely to be driven by changes in mitochondrial turnover or polarisation. AM treatment with the mitochondrial ATP synthase inhibitor Oligomycin significantly impairs efferocytosis (Supplemental Figure 3G, p=0.0312), demonstrating AM functional dependence on mitochondrial respiration. However, following ARDS EV treatment we observed increased AM mitochondrial respiration in the context of impaired efferocytosis, indicating that a shift in metabolic profile was not the primary driver of AM dysfunction.

**Figure 3:**
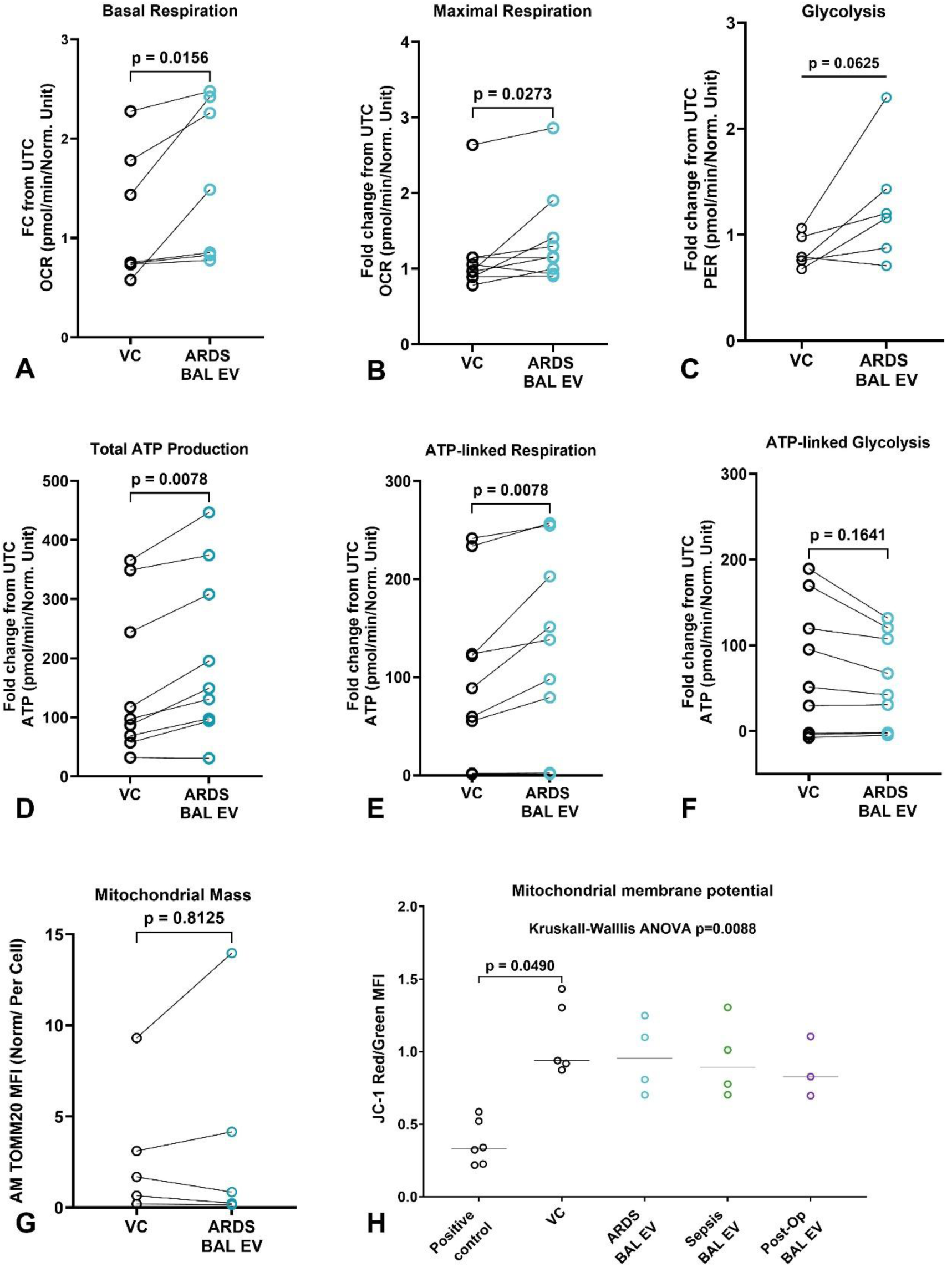
Pulmonary EVs on alveolar macrophage metabolic profile. Lobectomy AMs were treated with pooled EVs for 24 hours prior to assessment of metabolic profile. **A**: Impact of pooled ARDS BAL EV treatment on AM basal OCR (post glucose injection), n= 7. **B**: Impact of pooled ARDS BAL EV treatment on AM maximal OCR, n= 9. **C**: Impact of pooled ARDS BAL EV treatment on AM PER (post glucose injection, n=6. **D**: Impact of pooled ARDS BAL EV treatment on AM total ATP production, n=9. **E**: Impact of pooled ARDS BAL EV treatment on AM ATP-linked respiration, n=9. **F**: Impact of pooled ARDS BAL EV treatment on AM ATP-linked glycolysis, n=9. **G**: Impact of ARDS EV treatment on AM mitochondrial mass as assessed by TOMM20 expression, n=5. **H**: Impact of pooled BAL EVs from all patient groups on AM mitochondrial membrane depolarization as measured using JC-1, n=4 (Kruskal-Wallis test with Dunns multiple comparisons). AM: Alveolar macrophage, BAL: broncho-alveolar lavage, EV: extracellular vesicle, OCR: oxygen consumption rate, PER: proton efflux rate, TOMM20: Translocase of Outer Mitochondrial Membrane 20, UTC: Untreated control, VC: Vehicle control (saline).

### Impact of ARDS EVs on AM Autophagy

Due to the close interaction between metabolism and autophagy in modulating macrophage function (17), we assessed autophagosome and lysosome formation following ARDS patient EV treatment. Pooled ARDS patient BAL EV treatment reduced expression of AM autophagosome (LC3B-II, Figure 4A, p=0.001) and lysosome (LAMP, Figure 4B, p=0.0039) markers, suggestive of impaired autophagy machinery. Pooled ARDS patient NDBL EV treatment similarly reduced AM LAMP and LC3B-II expression (Supplemental Figure 4). These findings were further supported by chloroquine treatment, which revealed no increase in AM autophagosome accumulation, indicating suppressed autophagic flux in response to treatment with EVs from ARDS patients (Figure 4C). Thus, ARDS EV treatment impairs both AM autophagic flux and efferocytosis; reduced LC3 expression may lead to impaired LC3-associated efferocytosis (18).

**Figure 4:**
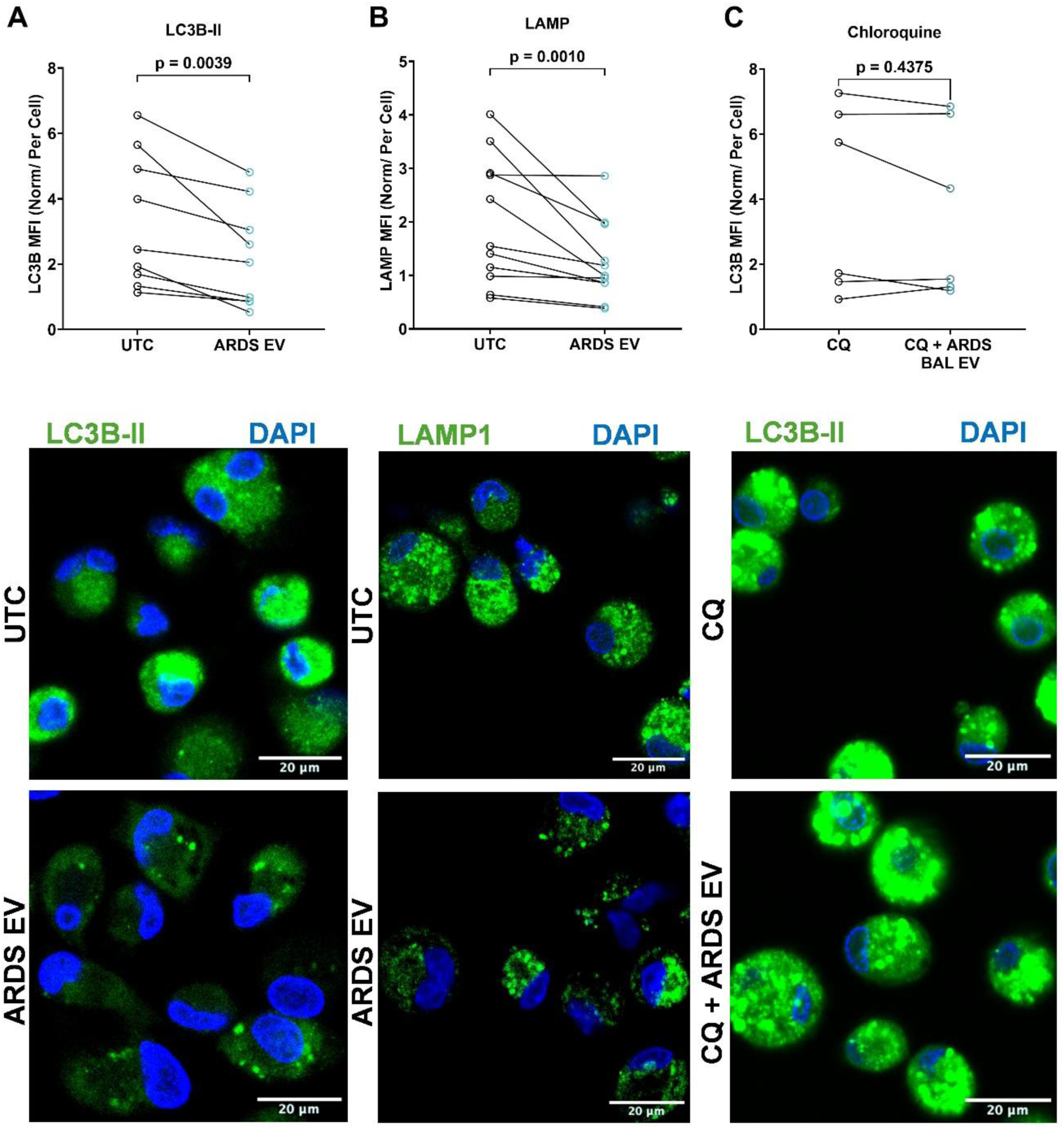
Pulmonary EVs on alveolar macrophage autophagy. Lobectomy AMs were treated with pooled EVs for 24 hours prior to assessment of autophagy. **A**: Impact of pooled ARDS BAL EV treatment on LC3B-II expression in AMs, n=9. Representative fluorescence microscopy image (x40): DAPI blue nuclear stain, LC3B-II = green. **B**: Impact of pooled ARDS BAL EV treatment on LAMP expression in AMs, n=11. Representative fluorescence microscopy image (x40): DAPI blue nuclear stain, LAMP1 = green. **C**: Impact of chloroquine treatment on AM LC3B-II expression, with or without prior treatment with pooled ARDS BAL EVs, n=6. Representative fluorescence microscopy image (x40): DAPI blue nuclear stain, LC3B-II = green. AM: Alveolar macrophage, BAL: broncho-alveolar lavage, CQ: Chloroquine, EV: extracellular vesicle, LAMP1: Lysosome-associated membrane protein-1, L3CB-II: Microtubule-associated proteins 1A/1B light chain 3B-II, UTC: Untreated Control.

### Characterisation of ARDS patient EV protein cargo

Proteomic analysis of individual BAL EV samples from sepsis patients with and without ARDS, and post-operative control patients was undertaken, with 7617 proteins identified (Figure 5A-B). Only 4 BAL EV samples from the postoperative group contained sufficient protein for analysis. 112 Proteins were upregulated and 132 proteins downregulated in sepsis patients with ARDS compared to sepsis patients without ARDS (Figure 5C). 231 Proteins were upregulated and 84 proteins downregulated in sepsis patients with ARDS compared to post-operative control patients (Figure 5D). IPA proteomic pathway analysis predominantly revealed significant downregulation of multiple pathways associated with immune cell function in the EVs of patients with sepsis-related ARDS compared to both sepsis patients without ARDS and postoperative controls (Figure 5E-F). These downregulated pathways associated with innate immune responses including ‘Binding and uptake of ligands associated with scavenger receptors’, ‘complement cascade’, ‘Fcγ receptor dependent phagocytosis’, and ‘cell surface interactions at the vascular wall’. This downregulation of multiple pathways associated with innate immune cell function demonstrates a potential role for EV protein cargo in mediating part of the functional impairment observed. However, this proteomic analysis of ARDS EVs did not identify a specific protein or single pathway which could represent a modifiable target to rescue AM function or autophagy flux.

**Figure 5:**
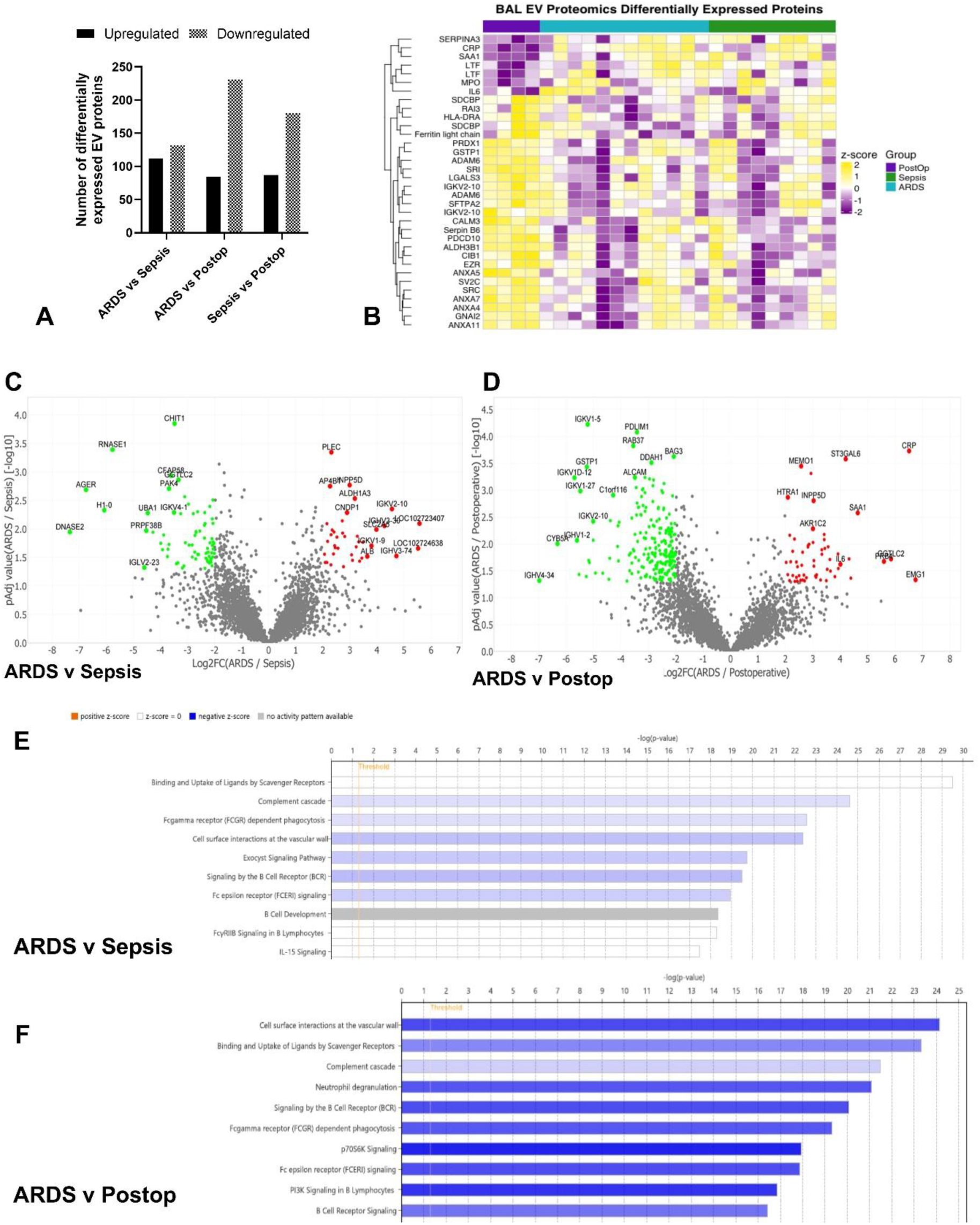
Proteomic analysis of BAL EVs. Proteomic analysis of individual BAL EV samples from sepsis patients with ARDS (n=12), sepsis patients without ARDS (n=9), and post-operative control patients (n=4) was undertaken, with 7617 proteins identified. **A**: Bar chart showing the number of differentially expressed BAL EV proteins between the three patient groups. **B**: Heatmap of differentially expressed BAL EV proteins between the three patient groups. **C**: Volcano plot of differentially expressed BAL EV proteins between sepsis patients with and without ARDS. **D**: Volcano plot of differentially expressed BAL EV proteins between sepsis patients with ARDS and Postoperative controls. **E**: IPA proteomic pathway analysis of differentially expressed BAL EV proteins between sepsis patients with and without ARDS. **F**: IPA proteomic pathway analysis of differentially expressed BAL EV proteins between sepsis patients with ARDS and postoperative controls.

### Characterisation of ARDS patient EV small RNA cargo

Small RNA transcriptomic analysis of individual BAL EV samples from post-operative control, sepsis with ARDS and sepsis without ARDS patients was undertaken (Supplemental Tables 1-3). 2731 mature microRNAs were identified (Supplemental Figure 5A). There were no differentially expressed microRNAs between sepsis patients with and without ARDS. 24 EV microRNAs were upregulated and 33 EV microRNAs were downregulated in sepsis patients with ARDS compared to post-operative control patients (Figure 6A). 22 EV microRNAs were upregulated and 18 EV microRNAs were downregulated in sepsis patients without ARDS compared to post-operative control patients (Figure 6B). Of these upregulated microRNAs, 15 were commonly upregulated across sepsis patients with and without ARDS. The most highly enriched EV microRNA in sepsis patients with and without ARDS compared to postoperative controls was miR-652-3p. There was no difference in total EV RNA isolated from equal volumes of BAL and NDBL fluid across all patient groups (Supplemental Figure 5B) RT-qPCR validated enrichment of BAL EV miR-652-3p in sepsis patients with ARDS compared to post-operative controls (Figure 6C, medians 0.00264 vs 0.0234, p=0.0185). However, RT-qPCR showed no difference in BAL EV miR-652-3p between sepsis patients without ARDS compared to post-operative controls (Figure 6C, p=0.136). RT-qPCR showed no difference in BAL EV content of the other 3 most highly enriched microRNAs identified from sequencing (miR-15a-5p, miR-193a-5p and miR-223-5p) between postoperative controls and sepsis patients with ARDS (Supplemental Figure 5C-E). RT-qPCR confirmed enrichment of miR-652-3p in the NDBL EVs of sepsis patients with and without ARDS compared to post-operative patient BAL EVs (Supplemental Figure 5F). NDBL EV subgroup analysis from sepsis patients with ARDS showed that CD14+ EVs had greater enrichment of mir-652-3p cargo compared to CD66+ (neutrophil-derived) and EpCAM (epithelial-derived) EVs (Figure 6D). Elevated NDBL EV miR-652-3p levels are associated with increased 90-day mortality in ARDS patients (Figure 6E, medians 9.20 vs 0.59 RQ, p=0.0295). Across all sepsis patient with and without ARDS, BAL EV miR-652-3p expression correlated negatively with oxygenation (Figure 6F, r=-0.698, p=0.0001). Diana-MirPath pathway analysis identified multiple KEGG pathways associated with miR-652-3p, including the Hippo signalling pathway (regulator of autophagy)(19) and regulation of actin cytoskeleton (required for efferocytosis) (Figure 6G). In the literature, microRNA-652-3p has been implicated in regulating macrophage lipid metabolism and pro-inflammatory effector functions; mRNA targets include TP53 which modulates autophagy (20, 21). Ingenuity Pathway Analysis (IPA, Qiagen) target filter analysis provides predicted microRNA–mRNA interactions from TargetScan, TarBase, miRecords and peer-reviewed literature. For miR-652-3p, IPA target filter confirmed TP53 as target and revealed novel targets including YWHAH, which activates PI3K / AKT pathways which promote efferocytosis (22, 23) (Supplementary Figure 5G). In summary, EV miR-652-3p was enriched in the BAL / NDBL of ARDS patients, associated with mortality and disease severity, and postulated to act via mRNA targets to suppress AM efferocytosis and autophagy flux.

**Figure 6:**
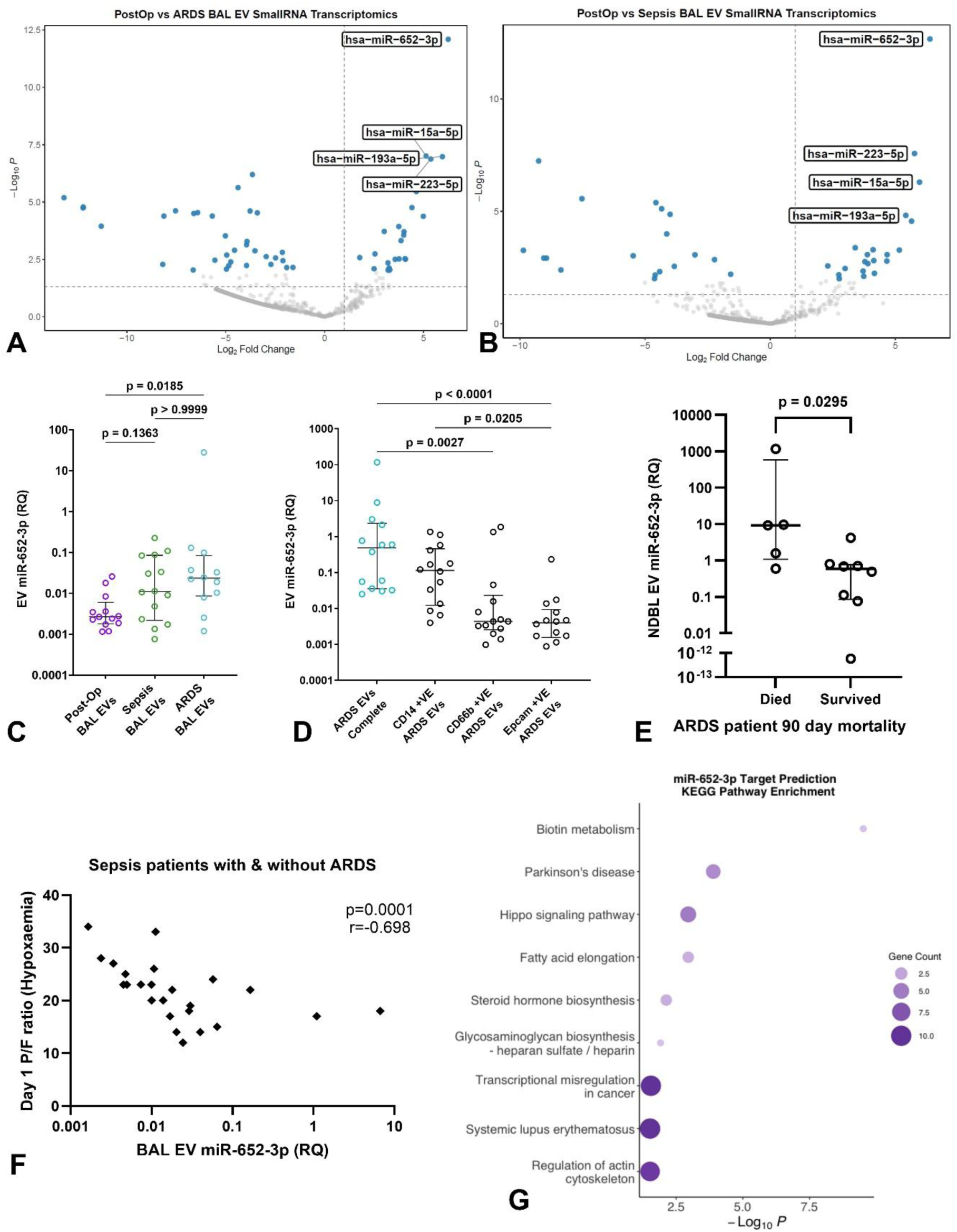
Small RNA transcriptomic analysis of BAL EVs. Small RNA transcriptomic analysis individual BAL EV samples from sepsis patients with ARDS (n=14), sepsis patients without ARDS (n=14), and post-operative control patients (n=13) was undertaken. **A**: Volcano plot of differentially expressed BAL EV microRNAs between post-operative controls and sepsis patients with ARDS (n≥13 in both groups, log2fold change >1.5, pAdj <0.05). **B A**: Volcano plot of differentially expressed BAL EV microRNAs between post-operative controls and sepsis patients without ARDS (n≥13 in both groups, log2fold change >1.5, pAdj <0.05). **C**: RT-qPCR validation of miR-652-3p expression within ARDS BAL EVs compared to post-operative controls (n≥13 in both groups, Kruskal-Wallis test with Dunns multiple comparisons). **D**: RT-qPCR analysis of miR-652-3p expression in isolated subpopulations (CD14^+^, CD66^+^, EpCAM^+^) of NDBL EVs from sepsis patients with ARDS (n=15, Friedman ANOVA with Dunns multiple comparisons). **E**: Association of NDBL EV miR-652-3p expression with 90-day mortality in sepsis patients with ARDS (n=15). **F**: Correlation of BAL EV miR-652-3p expression with P/F ratio on ICU admission across all sepsis patients, with and without ARDS (n=28). **G**: miR-652-3p target prediction KEGG pathway analysis from Diana-MirPath

### MicroRNA-652-3p mimic and antagomir studies

We set out to mechanistically determine the role of ARDS EV miR-652-3p in driving AM dysfunction. *Ex vivo* AMs from sepsis patients with ARDS have reduced expression of miR-652-3p targets p53 (Figure 7A, medians 0.000503 vs 0.000156, p=0.0299) and YWHAH (Figure 7B, medians 0.00116 vs 0.000161, p=0.0461) compared to AMs from lobectomy patients, supporting the role of miR-652-3p transfer in mediating AM dysfunction. YWHAH is a target of miR-652-3p and has been shown to protect against mitochondrial injury and promote autophagy. Direct transfection of lobectomy AMs with mimics of miR-652-3p significantly impaired efferocytosis (Figure 7C, p=0.0325), and co-treatment with antagomirs abrogates this functional impairment (Figure 7C, p>0.9999). MiR-652-3p mimic transfection inhibits AM LC3B-II expression (Figure 7D, mean of differences-1.24, p=0.0489). Transfection of pooled ARDS patient BAL EVs with antagomirs against miR-652-3p prior to AM treatment partially rescued the impairment of efferocytosis (Figure 7E, p=0.0143) and LC3B-II expression (Figure 7F, p=0.0437) compared to transfection with scramble antagomir. In summary, antisense oligonucleotides (antagomirs) against miR-652-3p can partially abrogate the suppressive impact of ARDS EVs on AM efferocytosis and autophagosome formation.

**Figure 7:**
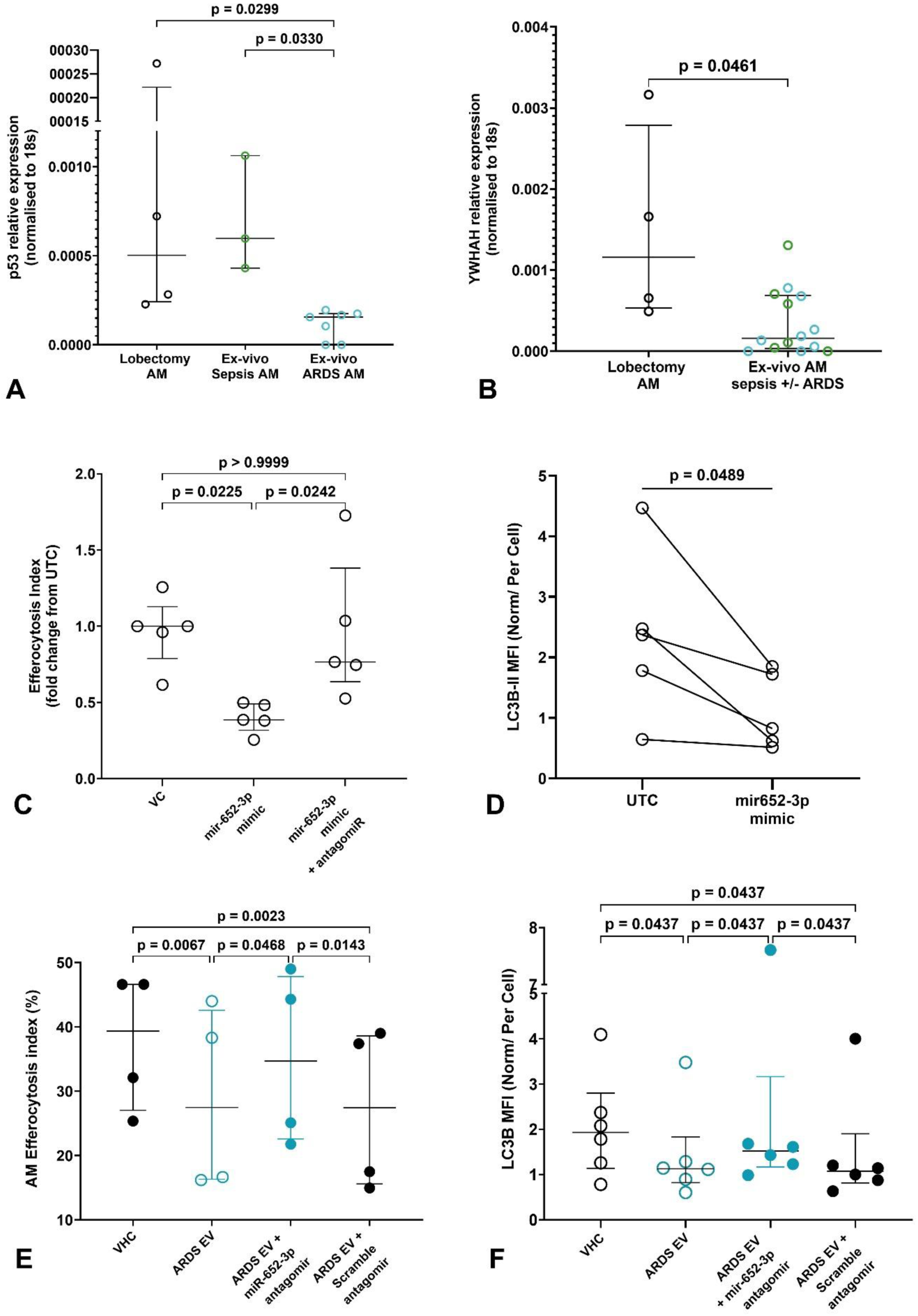
ARDS BAL EV miR-652-3p drives dysregulation of AM function and autophagy. Mechanistic studies were undertaken to determine the role of EV miR-652-3p in driving AM dysfunction **A**: RT-qPCR of p53 in *ex vivo* AMs from sepsis patients with ARDS, sepsis patients without ARDS, and from lobectomy patients, n≥3 all groups (Kruskal-Wallis test). **B**: RT-qPCR of YWHAH in *ex vivo* AMs from sepsis patients with and without ARDS, and from lobectomy patients, n≥4 all groups. **C**: Impact of lobectomy AM direct transfection with miR-652-3p mimics and inhibitors on efferocytosis, n=5 (Repeated Measures ANOVA with Dunns multiple comparisons tests). **D**: Impact of lobectomy AM direct transfection with miR-652-3p mimics on LC3B-II expression, n=5. **E**: Impact of ARDS EV transfection with miR-652-3p on lobectomy AM efferocytosis, n=4 (Repeated Measures ANOVA with Dunns multiple comparisons tests). **F**: Impact of ARDS EV transfection with miR-652-3p on lobectomy AM expression of LC3B-II, n=6 (Friedman ANOVA with Dunns multiple comparisons tests). AM: Alveolar macrophage, BAL: broncho-alveolar lavage, EV: extracellular vesicle, L3CB-II: Microtubule-associated proteins 1A/1B light chain 3B-II, UTC: Untreated Control, YWHAH: tyrosine 3-monooxygenase/tryptophan 5-monooxygenase activation protein eta.

## Discussion

In this study, we found that pulmonary EVs from patients with sepsis-related ARDS can induce dysregulation of AM efferocytosis, metabolic profile and autophagy. The same dysfunction is observed in *ex vivo* AMs from ARDS patients, and associated with increased mortality and duration of mechanical ventilation (4). Proteomic analysis revealed downregulation of multiple pathways associated with innate immune function, however no specific protein or single pathway was identified which could represent a modifiable target to rescue AM function. Small RNA sequencing revealed enrichment of miR-652-3p in the pulmonary EVs of sepsis patients (with and without ARDS). Pulmonary EV miR-652-3p expression was associated with increased 90-day mortality in ARDS patients, and directly correlated with the degree of hypoxemia. MicroRNA-652-3p was predominantly expressed within CD14^+^ (monocyte-derived) EVs, increased numbers of which have also been associated with higher mortality in ARDS (12). Mechanistic studies revealed that EV cargo miR-652-3p is a key driver of the major dysregulation of AM function and autophagy machinery observed in ARDS, indicating a role in pathogenesis. Inhibition of EV miR-652-3p resulted in partial restoration of AM function and autophagy. Thus, inhibition of pulmonary EV miR-652-3p may promote clearance of apoptotic neutrophils, attenuate inflammation and may offer a novel therapeutic strategy for ARDS patients.

We demonstrated that the EV component of BAL / NDBL fluid from ARDS patients predominantly drove the impairment in ARDS efferocytosis, as opposed to other components in these biofluids. We initially postulated that due to human AM reliance on mitochondrial respiration for ATP generation and effector functions including efferocytosis, that impaired efferocytosis would be associated with decreased mitochondrial bioenergetics. However, metabolic profiling revealed the converse, demonstrating an increase in AM ATP generation driven predominantly by increased mitochondrial respiration. Thus, changes in metabolic profile were not driving the impaired AM function observed; the specific EV component(s) inducing this increased mitochondrial activity remains to be determined. As AM mitochondrial activity is predominantly fuelled by fatty acid oxidation (24), EV lipidomic analysis may help to decipher these findings.

We found that ARDS pulmonary EV treatment significantly impairs autophagosome and lysosome formation in AMs. Autophagy is the process by which cells break down and recycle old and damaged organelles, maintaining cellular homeostasis. Impairment of macrophage autophagy has been associated with development of a pro-inflammatory phenotype and promotion of an inflammatory disease state (25, 26). Promotion of autophagy with rapamycin treatment has been shown to attenuate LPS-induced murine lung injury (27). A functional relationship between macrophage autophagy and efferocytosis has previously been identified (28). LC3 is critical for autophagosome biogenesis and maturation: on induction of autophagy, LC3-I is converted to LC3-II and recruited to autophagosomal membranes. LC3 is also required for LC3-associated phagocytosis / efferocytosis; this process involves recruitment of LC3 to phagosomes following engulfment of apoptotic cells, leading to rapid lysosomal fusion and degradation of contents (29, 30). In this way, the significant impairment in LC3B expression observed in AMs following ARDS EV treatment may directly contribute to impaired efferocytosis, linking autophagic machinery with AM function (17). Promotion of macrophage autophagy may enhance efferocytosis and thereby aid resolution of inflammation.

Previous studies and miRNA-mRNA target prediction models give some indication as to how miR-652-3p may mediate dysregulation of AM function and autophagy. Liu et al (20) showed that miR-652-3p inhibition in macrophages increased TP53 expression, and cholesterol efflux via increased expression of ABCA1 and ABCG1. Upregulation of ABCA1 in macrophages is associated with maintenance of lipid homeostasis and enabling recurrent efferocytosis (31, 32). TP53 encodes p53; nuclear p53 acts via damage-regulated autophagy modulator (DRAM) to induce autophagy (21, 33). Target prediction models also reveal tyrosine 3-monooxygenase/tryptophan 5-monooxygenase activation protein eta (YWHAH) as another target of miR-652-3p. YWHAH promotes autophagy via MAPK/ERK signalling (23). YWHAH also activates the PI3K signalling pathway, which promote efferocytosis (22, 34). Thus, miR-652-3p impairs the closely interlinked functions of AM efferocytosis and autophagy via multiple pathways.

This study shows that pulmonary EVs from BAL and NDBL fluid are bioactive when used to treat AMs, and are enriched in miR-652-3p. As collection of NDBL has lower risks compared to BAL in ARDS patients (35), and NDBL EV miR-652-3p is associated with greater 90-day mortality, this supports the prospect of utilising NDBL EVs and their microRNA cargo as potential biomarkers for ARDS. These findings also indicate that in ARDS patients, bioactive EVs containing miR-652-3p are present throughout the lower respiratory tract. Correlation of EV miR-652-3p levels with markers of ARDS severity (PaO_2_/FiO_2_ ratio) also highlights a potential role in prognosis. Studies investigating circulating EVs have shown differential expression of EV microRNAs between sepsis patients with and without ARDS (36), however no association with clinical outcomes was found and miR-652-3p was not enriched in the circulating EVs of ARDS patients. Pulmonary EV microRNAs likely represent a more relevant and sensitive biomarker for ARDS.

Antisense oligonucleotides (antagomirs) against miR-652-3p were able to partially rescue the impact of ARDS EVs on lobectomy AM efferocytosis and autophagy. As normal function was not fully restored, other EV components are also likely to play a contributing role in mediating AM dysfunction. EV proteomic analysis reveals downregulation of multiple pathways associated with innate immune function, however it was not possible to identify a single pathway or target for modulation. EV lipid cargo may also play a role in mediating macrophage function impairment, with further studies required to investigate this potential mechanism. However, the identification of EV miR-652-3p as a major contributor to AM dysfunction in ARDS offers a discrete therapeutic target.

MiR-652-3p is predominantly found within CD14^+^ pulmonary EVs, indicating that these EVs are released by the pro-inflammatory monocytes which infiltrate the alveolar space following an initial infection. These EVs are taken up by tissue-resident AMs distal to the original site of injury, which undergo modulation of their function and intracellular machinery that facilitates persistent inflammation. Therefore, pulmonary CD14^+^ EVs and/or their miR-652-3p cargo may represent a potential therapeutic target for patients with sepsis-related ARDS. Development of an inhaled antagomir therapy may abrogate the effects of EV miR-652-3p on AMs and potentially dampen excessive inflammation. Inhaled antisense oligonucleotides have successfully undergone phase 1 trials in patients with cystic fibrosis, indicating the feasibility of this approach (37, 38). Conjugation with surfactant proteins may aid uptake of antagomirs by AMs (39). Studies to evaluate off-target effects on alveolar epithelial cells would also be required, however inhibition of miR-652-3p can promote repair in endothelial cells (40). EVs and their cargo may represent novel therapeutic targets for ARDS, however, assessment of antagomir in both *in vivo* and human *ex vivo* lung models is first required.

Use of primary human AMs is a key strength of our study. This is particularly relevant as human AMs demonstrate divergence from monocyte-derived macrophages in their functional and metabolic responses to stimuli (41). There is also evidence to support elements of divergence between human and mouse AMs (42, 43). A limitation of our study is that pooling of EVs across patient groups was required to facilitate mechanistic investigation in lobectomy AMs, however this removes inter-individual variability for these experiments. Another limitation of our study is that other cargo components of EVs (e.g., lipids) may also be contributing to their biological effect on AMs and their role requires further elucidation. However, we have found that EV microRNAs play a major role and importantly offer a distinct therapeutic target in miR-652-3p.

In summary, pulmonary EVs (predominantly CD14^+^) from ARDS patients transfer miR-652-3p to alveolar macrophages. This transfer induces dysregulation of alveolar macrophage function and autophagy, which contributes to persistent inflammation and is associated with worse clinical outcomes including more hypoxemia and higher mortality. Thus, pulmonary EV miR-652-3p represents a novel therapeutic target for patients with sepsis-related ARDS.

## Methods

### Patient recruitment

The AM-ARDS study was conducted at the ICU of Queen Elizabeth Hospital Birmingham, U.K. from December 2016 to January 2019. The VESPER study was conducted at the same location from August 2023 to March 2026. For both studies, ethical approvals were obtained to recruit invasively ventilated adult sepsis patients, with and without ARDS (REC 16/WA/0169 and REC 22/LO/0872). Sepsis was defined according to Sepsis-3 criteria (44). For both studies, patients who fulfilled the Berlin criteria (45) within the previous 48 hours were classified as having ARDS. Exclusion criteria for both studies included imminent treatment withdrawal, prior steroid therapy, abnormal clotting, and clinically relevant immunosuppression. For both studies, pulmonary samples were collected within 48 hours of initiation of mechanical ventilation: BAL in the AM-ARDS study and non-directed bronchial lavage (NDBL) in the VESPER study.

In the VINDALOO trial (13), patients underwent postoperative bronchoscopy and BAL following oesophagectomy for oesophageal carcinoma (REC 12/WM/0092). Following unblinding of the study, those patients who received placebo (and did not later develop ARDS) were identified, and analysed as ventilated postoperative controls in this study.

The Birmingham Lung Tissue Study (REC 17/WM/0272) recruited adult patients scheduled to undergo lobectomy as part of their clinical treatment plan (predominantly for early stage lung cancer) at the Thoracic Surgery Unit in Queen Elizabeth Hospital Birmingham from September 2022 to March as previously described (5). Never-smoker or long-term ex-smoker (quit >5 years ago) patients with normal spirometry and no history of airways disease were recruited. Patients who had received immunotherapy or chemotherapy pre-operatively were not recruited. Human lung tissue samples distant from any tumour (if present), with no evidence of macroscopic pathology, and which were surplus to histopathological purposes, were immediately immersed in sterile 0.9% saline. Samples were transported on ice to the laboratory for isolation of alveolar macrophages.

### BAL and NDBL sample collection and processing

For BAL collection in the AM-ARDS study, an Olympus LF-TP fiberoptic scope (Olympus-Keymed, UK) was inserted through the patient’s endo-tracheal tube or tracheostomy tube, and the tip was wedged into a sub-segmental bronchus of the lingula or right middle lobe. Two 50 ml aliquots of sterile 0.9% saline at room temperature were instilled as a lavage, and the BAL fluid was aspirated. BAL collection in the Vindaloo study was undertaken in an identical way, immediately following esophagectomy. For NDBL collection in the VESPER study, a 14 Ch suction tube was inserted through the patient’s endo-tracheal tube or tracheostomy tube, and the tip placed 1cm above the carina. Two 50 ml aliquots of sterile 0.9% saline at room temperature were instilled as a lavage, and the NDBL fluid was aspirated. Both BAL and NDBL samples were immediately placed on ice and transported to the laboratory for identical processing. BAL and NDBL fluid was first filtered through sterile surgical gauze to remove mucus. Cell viability was assessed using trypan blue. Differential cell count was performed using cytospin and Diff-Quik labelling (Gentaur Europe, Kampenhout, Belgium). The filtered BAL and NDBL was then centrifuged at 560 g for 10 minutes at 4°C. Acellular BAL and NDBL supernatant was aspirated and stored in 1 ml aliquots at-80°C prior to EV isolation. The cell pellet was re-suspended in 10 ml of RPMI 1640 media (Sigma-Aldrich, UK) containing 10% Fetal Bovine Serum (FBS; Gibco, ThermoFisher, USA) prior to AM isolation.

### Extracellular Vesicle isolation from BAL and NDBL samples

Acellular BAL and NDBL samples were thawed once and ultracentrifuged at 100,000g (Beckman Coulter Optima MAX-XP ultracentrifuge) for 2 hours at 4°C. The EV-depleted supernatant was aspirated and stored at-80°C for further analysis. The EV pellet was resuspended in filtered PBS and ultracentrifuged again at 100,000g for 2 hours at 4°C. The EV pellets were then resuspended in 50µl PBS and pooled across patient groups.

### Human lung tissue processing

The airways of lung resection samples were immediately lavaged through with 500 – 2000 mls of sterile 0.9% saline (Baxter, UK) using a 14 gauge needle. The lavage fluid was centrifuged at 4°C and 560 g for 10 minutes and the supernatant discarded. The cell pellets were then pooled and re-suspended in 10 mls of RPMI 1640 media containing 10% FBS prior to AM isolation.

### Isolation of human alveolar macrophages

Re-suspended cell pellets from BAL and lung resection lavage were treated identically from this point onwards to isolate AMs as previously described (5). Mononuclear cells were separated by gradient centrifugation using Lymphoprep^TM^ (StemCell Technologies, Canada) as per manufacturer’s instructions. Isolated AMs were washed in PBS and purity assessed by cytospin. AMs were re-suspended in RPMI 1640 media supplemented with 10% FBS, 100 U/ml penicillin, 100 µg/ml streptomycin and 2 mM L-glutamine (Sigma-Aldrich) and plated at 2.5 x 10^5^ per well in a 24-well plate for functional assays, 8 x10^4^ per well for Seahorse and chamber-well seeding. AMs were cultured in an incubator at 37°C with 5% CO_2_ and media was changed after 24 hours to remove non-adherent cells. For AMs derived from lung tissue resections, flow cytometric staining with CD68 (Supplementary Table 5) was undertaken to confirm a pure population of AMs. If there was greater than 2% contamination of non-AM cells including interstitial macrophages, the sample was not utilised.

### Extracellular Vesicle Characterisation

#### Nanoparticle Tracking Analysis

EVs isolated by ultracentrifugation from patient BAL and NDBL samples were analysed by nanoparticle tracking analysis (NTA), using the NSPro (Malvern Panalytical, Malvern, UK), to determine size distribution and concentration using NanoSight NS Xplorer software (Malvern Panalytical, Malvern, UK). Automated detection threshold and focus were utilised. For each sample, 5 videos capturing 400 tracks were taken at 25 °C with a syringe pump speed of 3 μL/min.

#### Single particle interferometric reflectance image sensing (Exoview)

The EV phenotype in patient NDBL samples was characterised using the ExoView R100 reader (Unchained Labs, USA) to analyse size, concentration, and EV surface markers via Leprechaun Exosome Human Tetraspanin Kits (Unchained Labs, USA) as previously described (12). NDBL samples were diluted at 1:20 with incubation solution and incubated with tetraspanin chips for 16 hours to allow EV binding. Chips were then stained with a fluorescent antibody cocktail for 1 h at room temperature in the dark to characterize EV surface markers (Supplementary Table 5). Chips were then washed thrice and imaged using the ExoView R100 reader with ExoViewer 3.14 software and analysed using ExoView Analyser 3.0. Fluorescence gating was based on mouse IgG control, and sizing thresholds were set for a diameter of 50 - 200 nm.

#### Transmission Electron Microscopy

BAL EV pellets were incubated in 4% paraformaldehyde (PFA) at room temperature for 30 minutes. EV pellets were washed three times with PBS then resuspended in 20µl PBS prior to loading on Carbon-coated grids. The grids were dried then fixed with 2% glutaraldehyde. EVs were visualised on a TEM microscope (JEOL, JEM 1400Plus, Japan).

### Extracellular Vesicle small RNA transcriptomics

EVs were isolated from 1ml of BAL fluid using the ultracentrifugation protocol described above. Total RNA was extracted from the EV pellet using Trizol according to manufacturer’s instructions (ThermoFisher, UK). RNA quantity and purity was determined by Nanodrop One (ThermoFisher, UK). Library preparation and RNA-sequencing were performed by Beijing Genomics Institute (BGI, Hong Kong, China) using the DNBSEQ G400 platform to obtain 49-nt single-end sequencing reads. miRNA expression (reads) and differential expression were assessed using miRge3 (46) using the following command line:

miRge3.0-s <list of comma separated FASTQ files to be processed>-lib <pathway to hg38 indexed genome>-db miRBase-o <output directory>-dex-mdt <pathway to list of csv file of samples for differential expression testing>

The miRge3 programme aligns FASTQ data to the human hg38 reference genome and quantifies miRNA expression using miRbase. Differential expression was performed using DESeq2 (47). Ingenuity Pathway Analysis microRNA target filter was used to identify mRNA targets for upregulated miRNAs (IPA, Qiagen, UK).

### Real-Time quantitative Polymerase Chain Reaction (RT-qPCR)

RT-qPCR of EV microRNA was undertaken on both complete EV populations and subpopulations. Magnetic beads (EasySep™ EV PE positive isolation kit, StemCell Technologies) were used to isolate CD14+, CD66b+ and EpCAM+ EV subpopulations via PE conjugated antibodies (Supplementary Table 5) as per manufacturer’s instructions. Trizol reagent (ThermoFisher Scientific) was used to isolate total RNA from EV total/ sub-populations as per manufacturer’s instructions. RNA was quantified by a NanoDrop One Spectrophotometer (Thermofisher Scientific). 10ng RNA was reverse transcribed to cDNA using a Taqman Advanced microRNA cDNA synthesis kit (ThermoFisher Scientific). Following this, RT-qPCR was undertaken in triplicate using a Quantinova RT-PCR kit (Qiagen) and Taqman Advanced microRNA assays (ThermoFisher, Supplementary Table 4) on a CFX Opus Real-Time PCR Detection System (Biorad). EV miRNA levels were normalised to miR-28-5p, a uniformly expressed internal control identified via Normfinder software, using the Delta-Delta Ct method.

For quantification of cellular mRNA expression, total RNA was isolated using Trizol reagent. One-Step Quantifast Probe RT-PCR Kits (Qiagen, Hilden, Germany) were used with Taqman gene expression assays (ThermoFisher, Supplementary table 4). The High-Capacity cDNA Reverse Transcription Kit (Applied Biosystems) and PowerUP SYBR Green PCR kit (ThermoFisher) were used for qSTAR qPCR gene expression assays (OriGene, Supplementary table 4).

PCR conditions were used as per the recommendation of the manufacturer. Triplicate data were analysed using CFX Maestro software (BioRad). Relative quantification of target gene mRNA was calculated relative to expression of 18s endogenous control gene.

### Extracellular Vesicle Electroporation

Pooled EVs were electroporated with RNAse or Proteinase K as previously described (48). EVs were resuspended in 100µl OptiMEM (ThermoFisher) containing 5µg/ml RNAse (Merck) or 2µg/ml Proteinase K (ThermoFisher Scientific) and loaded into a Nucleocuvette® (Lonza). The cuvette was placed into the 4D-Nucleofector® X Unit (Lonza) and the DN-100 electroporation protocol was started. EV membrane integrity post-electroporation was determined using calcein staining on the Cytoflex Nano flow cytometer (Beckman Coulter). Following electroporation, EVs were incubated for 1 hour at 37°C for membrane recovery. EVs were then isolated from the electroporation reaction mixture via ultracentrifugation (Beckman Coulter Optima MAX-XP ultracentrifuge) at 100,000g and 4°C for 2 hours.

### Alveolar Macrophage Treatments

Regarding *in vitro* EV treatment of AMs, we treated 250,000 AMs with the equivalent number of EVs present in 250µl of pooled BAL or NDBL fluid. This was based on previous BAL treatment studies (6) to more closely reflect the *in vivo* alveolar environment in different patient groups. Functional and metabolic readouts were measured 24 hours after EV treatment. Other treatments included 50 ng/ml interferon-γ (IFN-γ, Peprotech) and 1 μg/ml ultrapure lipopolysaccharide (LPS, Invitrogen). AMs were also directly transfected with synthetic mimics and antagomirs of hsa-miR-652-3p as described above.

### Alveolar macrophage efferocytosis assay

This protocol was performed as previously described (6). Briefly, neutrophils were isolated from the blood of healthy volunteers using Percoll density centrifugation. Neutrophils were suspended in a 5μM solution of CellTracker^TM^ Deep Red fluorescent dye (ThermoFisher) at 4×10^6^/ml, then incubated for 30 minutes at 37°C. Stained neutrophils were centrifuged at 1500g for 5 minutes then incubated in serum-free RPMI at 37°C and 5% CO_2_ for 24 hours to induce apoptosis. As negative control, 5μg/ml Cytochalasin D (Sigma-Aldrich) was added to AMs for 30 minutes to inhibit actin filament polymerization. Stained apoptotic neutrophils were added to AMs at a 4:1 ratio prior to incubation for 2 hours at 37°C. Media was removed and wells washed twice with ice-cold PBS. Cells were harvested prior to acquisition using an MACSQuant 10 flow cytometer (Miltenyi Biotec). Background fluorescence from negative controls was subtracted from the percentage of APC+ AMs in other experimental conditions, to give a corrected net efferocytosis index representative of neutrophil engulfment.

### Alveolar Macrophage Metabolic activity assays

Real time analysis of the metabolic activity of AMs treated with pooled EVs was conducted using XFe96 Seahorse analyser (Agilent). Oxidative phosphorylation and glycolytic flux were determined using an adapted combined Mito-Glyco stress test assay, as previously described (49). The uncoupler FCCP was substituted for BAM-15 as this compound is less toxic and better tolerated by AMs. 80,000 cells were seeded per well with ≥6 replicate wells per condition. After seeding, cells were incubated at room temperature for 45 mins before transfer to 37°C and 5% CO2 incubation. After an initial overnight incubation, EV treatments were added for 24 hours. 1 hour prior to XF assay; cell culture media was washed off the AMs and replaced by Agilent Seahorse XF RMPI Medium (Agilent Technologies) at pH 7.4 containing Sodium Pyruvate (100nm) and L-Glutamine (200nm). Cells were then incubated for 1 hour (37°C, 0% CO_2_) prior to running the assay. The following assay injections and final concentrations were then used: (i) 10nM glucose, (ii) 1μM oligomycin, (iii) 2µM N5,N6-bis(2-Fluorophenyl)-oxadiazolo[4-b]pyrazine-5,6-diamine (BAM15) + 1nM sodium pyruvate, and (iv) 100nM rotenone + 1μM antimycin A + 20mM 2-deoxyglucose. To normalise for cell number variability and account for the presence of dead cells immediately after XF assay, Calcein-AM dye was used. On completion of the assay supernatant was removed from all wells, then Calcein-AM (1uM) dye was added to all cells and incubated for 45 minutes at 37°C. Excitation and emission 490nm/515nm were then read using a plate reader (Synergy 2, Bio-Tek, USA).

Wave software (Agilent technologies) was used for the initial analysis of XF assay results. Normalised OCR pmol/min and PER pmol/min readings were used for metabolic parameter calculations. To allow for the comparison of ATP generated at baseline by both types of metabolism, analysis was performed using the quantification of intracellular rates of ATP production using flux rates as reference (50). The conversion of amount of ATP made providing the OCR/ PER flux rates across basal measurements and during the glucose injection was calculated.

### Immunocytochemistry

Cells were cultured in chamber-wells at 80,000 cells per well prior to imaging (ibdi, Thistle Scientific, UK). Cells were fixed in 2% paraformaldehyde for 15 minutes, rinsed in PBS, then permeabilised and blocked (10% FBS, 0.1% TritonX-100 [Sigma-Aldrich]). Cells were then incubated overnight at 4°C with primary antibodies (Supplementary Table 5) in 10% FBS and 0.05% Tween-20 [Sigma-Aldrich], PBS. Following incubation, cells were probed with fluorophore-conjugated secondary antibodies (Supplementary Table 5) at room temperature for 90 minutes in the dark. The secondary antibody was washed off and cells were stained with DAPI (Merck, UK) for 5 minutes, at room temperature in the dark. Confocal images were acquired under constant photomultiplier settings (LSM 880 with Airyscan, Zeiss, UK) and processed using Python via the Jupyter platform software. Images were selected from the same cardinal points of the chamber well; at least 50 cells across 3 biological replicates were analysed. To assess autophagic flux, after ARDS BAL EV treatment, cells were treated with 50 μM of chloroquine for two hours before fixing (ThermoFisher Scientific, UK).

### Mitochondrial membrane potential assay

AM mitochondrial membrane potential was assessed via JC-1 staining (Mitochondrial Membrane Potential Assay Kit, Abcam) as per manufacturer’s instructions. Following EV treatment for 24 hours, AMs were cultured 0.5 μg/mL JC-1 dye for 30 minutes at 37°C prior to assessment of the red: green fluorescence ratio via flow cytometry (MACSQuant 10, Miltenyi Biotec). Positive controls were treated with Carbonyl cyanide 4-(trifluoromethoxy)phenylhydrazone (FCCP, 100 μM; Abcam) to uncouple mitochondria.

### MicroRNA mimic and antagomir transfection of EVs and AMs

Lipofectamine 2000 (ThermoFisher Scientific) was used to both transfect AMs directly, and transfect pooled EVs prior to AM treatment, with synthetic mimics or inhibitors of hsa-miR-652-3p and negative controls (Qiagen, UK) as per manufacturer’s instructions and as previously described (51). For direct mimic and inhibitor (antagomir) transfection of AMs, 5nM mimic and 15nM inhibitor (antagomir) concentrations were used. For direct transfection of EVs with microRNA inhibitor (antagomir), 200nM concentration was used. Following transfection, EVs were isolated from the lipofectamine reaction mixture via ultracentrifugation 100,000g at 4°C for 2 hours, then used to treat AMs.

hscmic sequence: 5’-AAUGGCGCCACUAGGGUUGUG-3’ (catalogue no. MSY0003322).

has-miR-652-3p power inhibitor sequence: 5’-ACAACCCTAGTGGCGCC-3’ (catalogue no. 339131 Y104101747-DDA).

Negative control A (scramble) miRNA power inhibitor control sequence: 5’-TAACACGTCTATACGCCCA-3’ (catalogue no. 339136 Y100199006-DDA).

### EV proteomics

Following ultracentrifugation, BAL EV pellets were resuspended in 50µl PBS. Untargeted Data Independent Analysis (DIA) proteomics were undertaken by Beijing Genomics Institute (BGI, Hong Kong, China) via liquid chromatography mass spectrometry.

#### Sample preparation

EV pellets were mixed with sodium dodecyl sulfate solution for lysis. Samples were reduced by adding 10nM dithiothreitol and heated at 37°C for 30 minutes, then alkylated with 55nM iodoacetamide in the dark for 45 minutes. Protein samples were then dissolved in 0.5M triethylammonium bicarbonate, centrifuged at 12,000g, then digested with trypsin. The collected peptide fractions were then freeze-dried.

#### Protein quantification

Dried peptide samples were reconstituted with mobile phase A (2% acetonitrile, 0.1% formic acid), centrifuged at 20,000g for 10 minutes. Peptides were separated using a Thermo UltiMate 3000 UHPLC liquid chromatograph. The sample was first enriched in the trap column and desalted, and then transferred to a tandem self-packed C18 column (150μm internal diameter, 1.8μm column size, 35cm column length), and separated at a flow rate of 500nL/min by the following gradient: 0-5 minutes, 5% mobile phase B (98% acetonitrile, 0.1% formic acid); 5-120 minutes, mobile phase B linearly increased from 5% to 25%; 120-160 minutes, mobile phase B rose from 25% to 35%; 160-170 minutes, mobile phase B rose from 35% to 80%; 170-175 minutes, 80% mobile phase B; 175-180 minutes, 5% mobile phase B.

For data-independent acquisition (DIA) analysis, LC separated peptides were ionized by nano electrospray ionization and injected to tandem mass spectrometer Q-Exactive HFX Hybrid Quadrupole-Orbitrap Mass Spectrometer (ThermoFisher Scientific, San Jose, CA) with DIA (data-independent acquisition) detection mode. The main settings were: ion source voltage 1.9kV; MS scan range 400∼1,250m/z; MS resolution 120,000, MIT 50ms; 400∼1,250m/z was equally divided to 45 continuous windows MS/MS scan. MS/MS collision type HCD, MIT was auto mode. Fragment ions were scanned in Orbitrap, MS/MS resolution 30,000, collision energy was distributed mode: 22.5, 25, 27.5, Automatic gain control was 1E6.

For DIA data, mProphet algorithm was used to complete analytical quality control. The acquired data were searched against the human proteome sequence database UniProt (https://www.uniprot.org/uniprot/). Based on the quantitative results, the differential proteins between comparison groups were found. The DIA data was analysed using the indexed retention time (iRT) peptides for retention time calibration. Then, based on the target-decoy model applicable to SWATH-MS, false positive control was performed with FDR 1%, to obtain significant quantitative results. This procedure extracted peak areas and calculated protein quantitation values. According to set comparison groups, the multiples of differences in the proteins in each comparison group were calculated. Protein intensity values were normalized by Log2 transformation. The mass spectrometry proteomics data have been deposited to the ProteomeXchange Consortium via the PRIDE repository with the dataset identifier PXD079596. Significantly differentially abundant proteins (filtered as log2fold >2 and pAdj <0.05) were uploaded to Ingenuity® Pathway Analysis (IPA®) software, Qiagen (Hilden, Germany) for further pathway analysis.

### Alveolar Macrophage Apoptosis and Viability assay

An Apoptosis / Dead Cell kit (Thermofisher Scientific) was used to assess AM viability and apoptosis. This kit incorporated FITC-Annexin V and PI stains which were used as per manufacturer’s instructions and measured via flow cytometry (MACSQuant 10, Miltenyi Biotec).

### Alveolar Macrophage Phagocytosis assay

Alveolar macrophage flow cytometric phagocytosis assays were performed using heat killed *S. pnemoniae* labelled with cell tracker Far Red. The TIGR4 (serotype 4) strain of *S. Pneumoniae* was suspended in a 5μM solution of AlexaFluor™ 647 Succinimidyl Ester (ThermoFisher) at 50million CFU /ml /ml, then incubated for 45 minutes at 37°C. Bacteria were then heat killed at 65°C for 2 hours. *S. Pneumoniae* were opsonised in 10% human sera one hour prior to experiment (SigmaAldrich), before centrifugation at 1200g, resuspension in PBS and sonication. As negative control, 5μg/ml Cytochalasin D (Sigma-Aldrich) was added to AMs for 30 minutes to inhibit actin filament polymerization. Stained heat-killed *S. pnemoniae* were added to AMs at a 50:1 ratio prior to incubation for 6 hours at 37°C. Media was removed and wells washed twice with ice-cold PBS. Cells were harvested prior to acquisition using an MACSQuant 10 flow cytometer (Miltenyi Biotec). Background fluorescence from negative controls was subtracted from the percentage of APC+ AMs in other experimental conditions, to give a corrected net phagocytosis index representative of bacterial engulfment.

## Statistical analyses

Statistical analysis was performed using GraphPad Prism v.10 software. Normality of data was assessed using the D’Agostino & Pearson test. Two-tailed p-values of ≤0.05 were considered as significant. Results from parametric data are shown as mean and standard deviation. Results from non-parametric data are shown as median and interquartile range. Differences between continuously distributed non-parametric data were assessed using Mann-Whitney tests. Differences between non-parametric paired data were assessed using the Wilcoxon matched-pairs signed rank test. Differences between paired parametric data were assessed using a paired t-test. Linear associations between non-parametric data sets were assessed using Spearman’s correlation coefficient. Differences between three or more non-parametric data sets were assessed using the Kruskal-Wallis one-way analysis of variance (ANOVA) and Dunn’s multiple comparison tests. Differences between three or more paired non-parametric data sets were assessed using the Friedman one-way ANOVA and Dunn’s multiple comparison tests. Differences between three or more paired parametric data sets were assessed using the repeated-measures ANOVA and Dunn’s multiple comparison tests.

## Study Approval

Ethical approvals were given by the UK Health Research Authority (HRA) Research Ethics Committee (REC) for patient recruitment for the following studies: AM-ARDS (16/WA/0169), VESPER (22/LO/0872), VINDALOO (12/WM/0092) and the Birmingham Lung Tissue Study (17/WM/0272). For the AM-ARDS and VESPER studies, patients were unable to give informed consent due to alterations in conscious level caused by therapeutic sedation. Therefore, their next of kin or a legal representative were requested to give assent for the patient to be recruited into the study. in accordance with the UK Mental Capacity Act (2005). For the Vindaloo and Birmingham Lung Tissue Studies, all patients gave informed written consent prior to participation.

## Author contributions

KLS and CM contributed to sample processing, laboratory data generation and analysis. JP provided technical expertise on EV characterisation. KS, CM, EJ, CC, XJ, SQ, LEC, BN, DP and RYM contributed to patient recruitment, sample collection, and clinical data collection. JRH provided technical expertise on assessment of autophagy. ML contributed to small RNA transcriptomic data analysis. MAM, DRT, DP, and AS critically commented on the study and revised the manuscript. KLS and RYM designed the study, analysed the data and wrote the manuscript. All authors critically reviewed the manuscript and approved the final draft for submission.

## Funding support

RYM reports Medical Research Council funding: (grant no. MR/N021185/1 and MR/X000338/1). AS, DT, DP report funding by Asthma+Lung UK (grant no. MCFPHD20F/2), Efficacy and Mechanism Evaluation (grant no. NIHR131600), Health Technology Assessment (grant no. NIHR129593), and Medical Research Council (grant no. MR/S002782/1, UKRI1465).

## Data Availability

All data produced in the present study are available upon reasonable request to the corresponding author

## Acknowledgements

We would like to thank the department of Thoracic surgery at University Hospitals Birmingham NHS Trust and tissue co-coordinators Nawaal Kiwia and Emmanuel Abimbola for their support with the lung tissue study. We would like to thank Leo da Cruz for his assistance with the graphical abstract. We would like to thank all patients, relatives and carers involved in this study. We would like to thank Malvern Panalytical for their assistance with nanoparticle tracking analysis.

## Notes

### Competing Interest Statement

MAM reports grants from Roche Genentech and Quantum Therapeutics; consulting fees from Healios Pharmaceuticals and Marck Pharmaceuticals. All other authors declare that no conflicts of interest exist.

