## Supplementary Information for "Pulmonary extracellular vesicles drive alveolar macrophage dysfunction via microRNA transfer in Acute Respiratory Distress Syndrome"

### SUPPLEMENTARY FIGURES

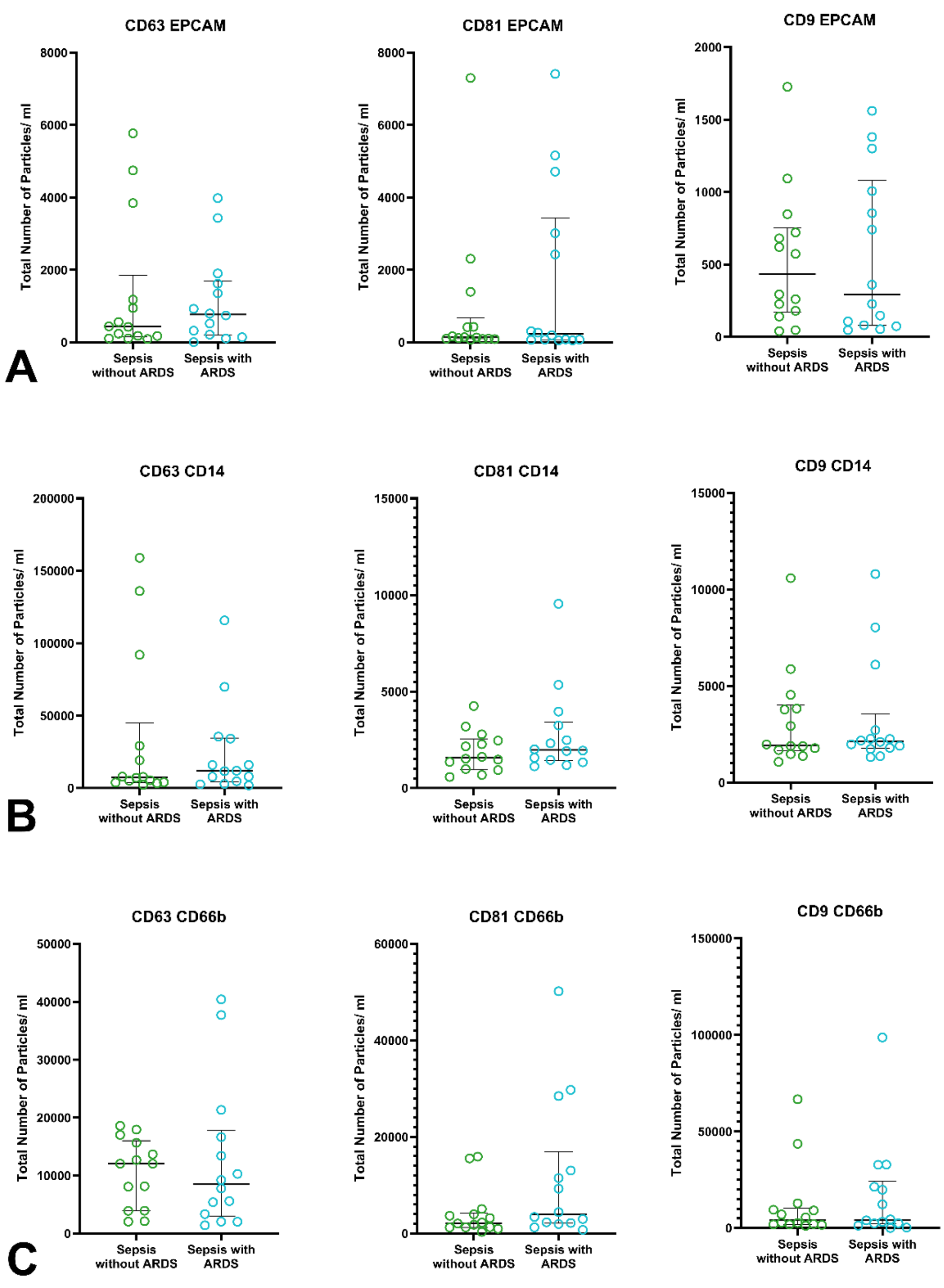


**Supplemental Figure 1. A:** Concentration of EVs following isolation from NDBL and pooling across patient groups, as measured by NTA. **B-D:** Exoview characterisation of NDBL EVs from sepsis patients with and without ARDS, n≥12. EVs were assayed for expression of surface markers including tetraspanins CD63, CD81 and C9 via antibody capture. Cell-specific surface markers were also assayed including EpCAM, CD14, and CD66b via antibody detection.

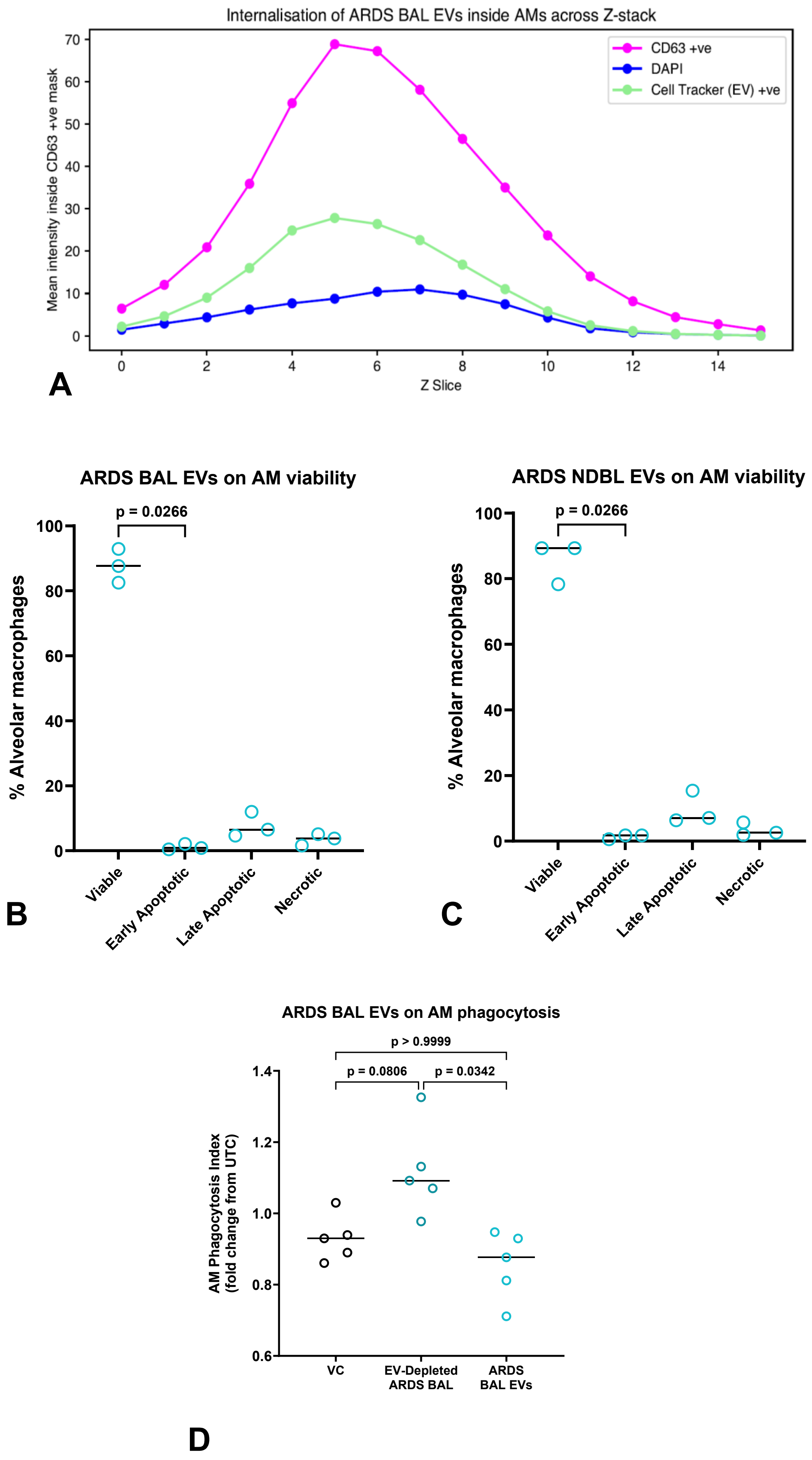
Internalisation of ARDS BAL EVs inside AMs across Z-stack
Mean intensity inside CD63 +ve mask
Z Slice
CD63 +ve
DAPI
Cell Tracker (EV) +ve
0 10 20 30 40 50 60 70
0 2 4 6 8 10 12 14
A
ARDS BAL EVs on AM viability
p = 0.0266
% Alveolar macrophages
0 20 40 60 80 100
Viable
Early Apoptotic
Late Apoptotic
Necrotic
B
ARDS NDBL EVs on AM viability
p = 0.0266
% Alveolar macrophages
0 20 40 60 80 100
Viable
Early Apoptotic
Late Apoptotic
Necrotic
C
ARDS BAL EVs on AM phagocytosis
p > 0.9999
p = 0.0806
p = 0.0342
AM Phagocytosis Index
(fold change from UTC)
0.6 0.8 1.0 1.2 1.4
VC
EV-Depleted ARDS BAL
ARDS BAL EVs
D

**Supplementary Figure 2. A:** Representative z-stack histogram demonstrating internalisation of celltracker green labelled EVs within DAPI and CD63 labelled AMs. **B:** Impact of ARDS patient pooled BAL EV treatment on AM viability and apoptosis, n=3. **C**: Impact of ARDS patient pooled NDBL EV treatment on AM viability and apoptosis, n=3. **D:** Impact of ARDS patient pooled BAL EV treatment on AM bacterial phagocytosis, n=5. AM: Alveolar macrophage, BAL: broncho-alveolar lavage, EV: extracellular vesicle, NDBL: non-directed bronchial lavage, UTC: Untreated control, VC: Vehicle control (50% saline).

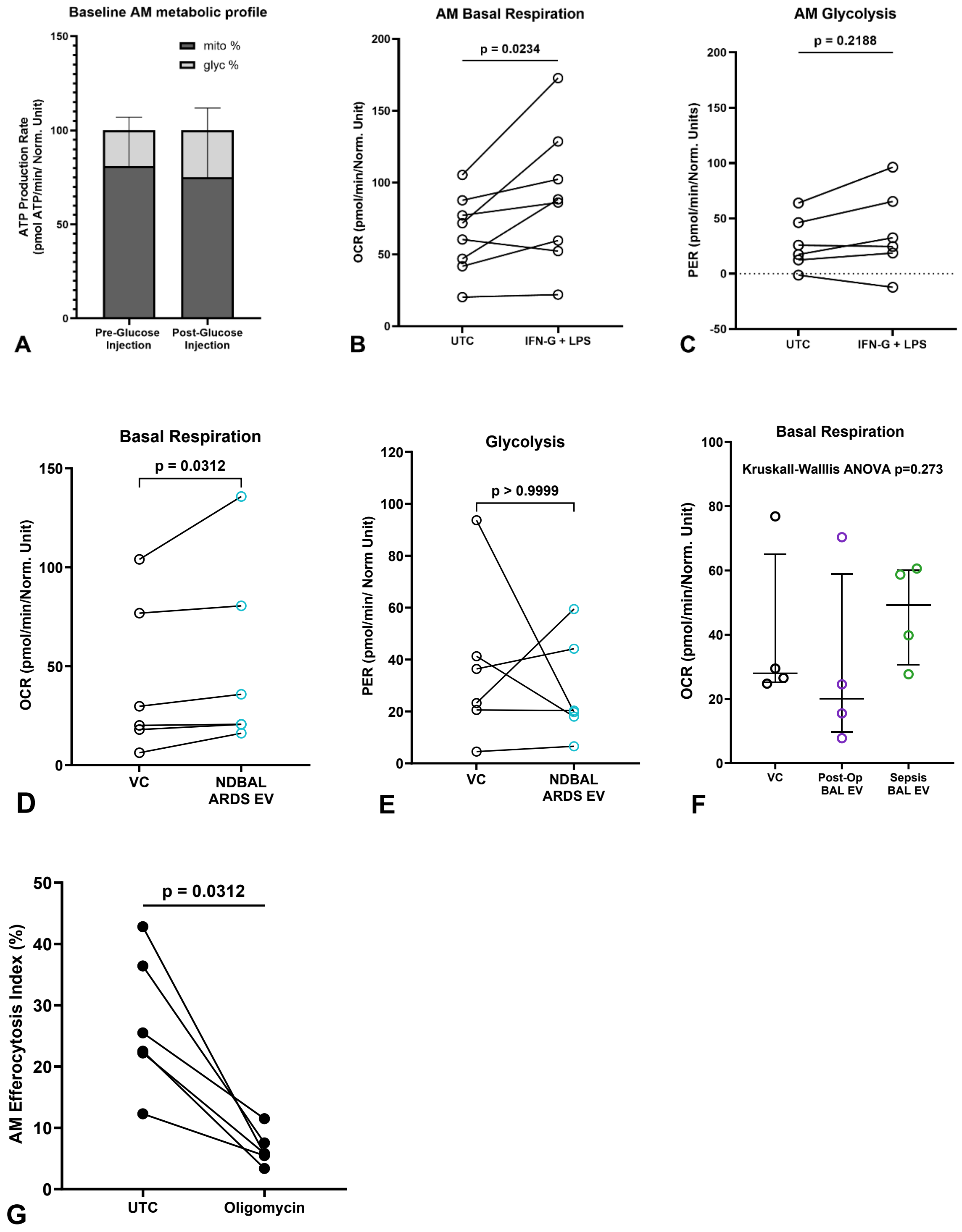


**Supplementary Figure 3. A:** Baseline AM metabolic profiling reveals a reliance on mitochondrial respiration to generate ATP and minimal glycolytic activity, both in the presence and absence of glucose. **B:** Stimulation of AMs with pro-inflammatory mediators (LPS and IFNγ) upregulated the mitochondrial activity (OCR), n=8. **C:**

Stimulation of AMs with pro-inflammatory mediators (LPS and IFNγ) had no impact on glycolysis (PER), n=6. **D-E:** Pooled NDBL EVs from sepsis patients with ARDS increased AM mitochondrial activity (OCR) but had no impact on glycolysis (PER), n=6. **F:** Impact of pooled BAL EVs from sepsis patients without ARDS and postoperative control patients on AM basal respiration, n=4. **G:** Impact of 30mins Oligomycin treatment on AM efferocytosis, n=6. AM: Alveolar macrophage, EV: extracellular vesicle, IFNγ: Interferon-γ, LPS: lipopolysaccharide, NDBL: non-directed bronchial lavage, OCR: oxygen consumption rate, PER: proton efflux rate UTC: Untreated control, VC: Vehicle control (saline).

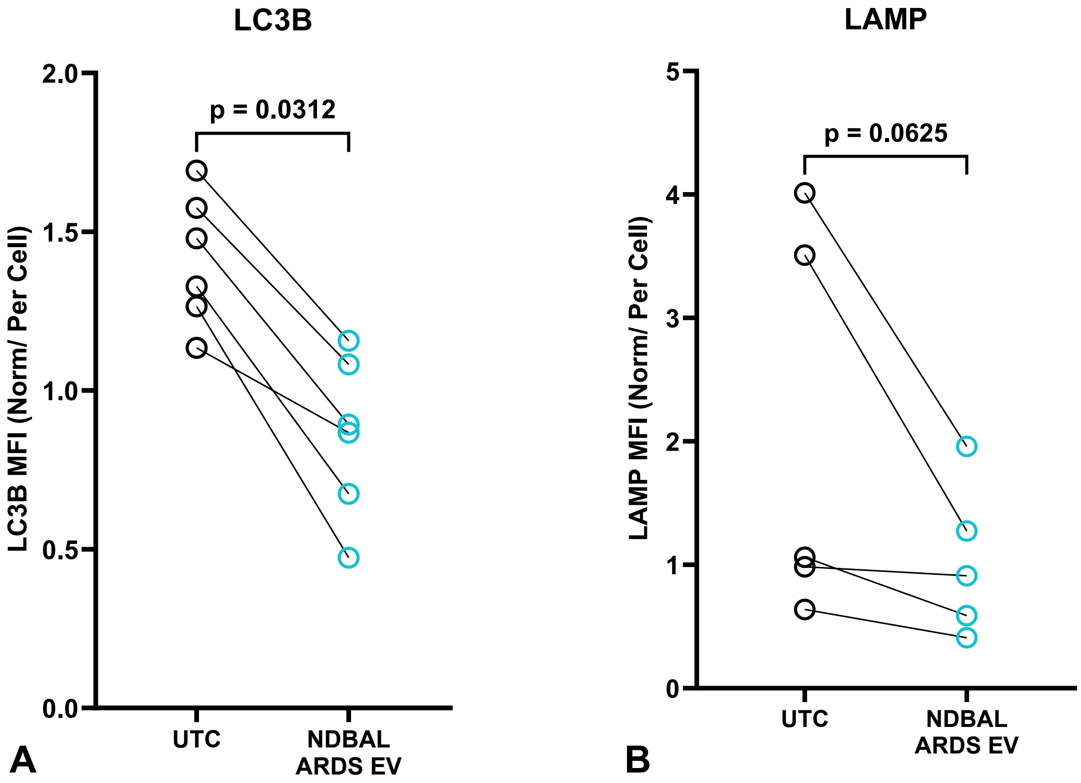


**Supplementary Figure 4. A:** Pooled ARDS patient NDBL EV treatment reduced AM LC3B-II expression, n=6. **B**: Pooled ARDS patient NDBL EV treatment showed a trend towards reduced AM LAMP expression, however this did not reach significance, n=5. AM: Alveolar macrophage, EV: extracellular vesicle, LAMP: Lysosomal-associated membrane protein 1, LC3B-II: Microtubule-associated proteins 1A/1B light chain 3B-II, NDBL: non-directed bronchial lavage, UTC: Untreated control.

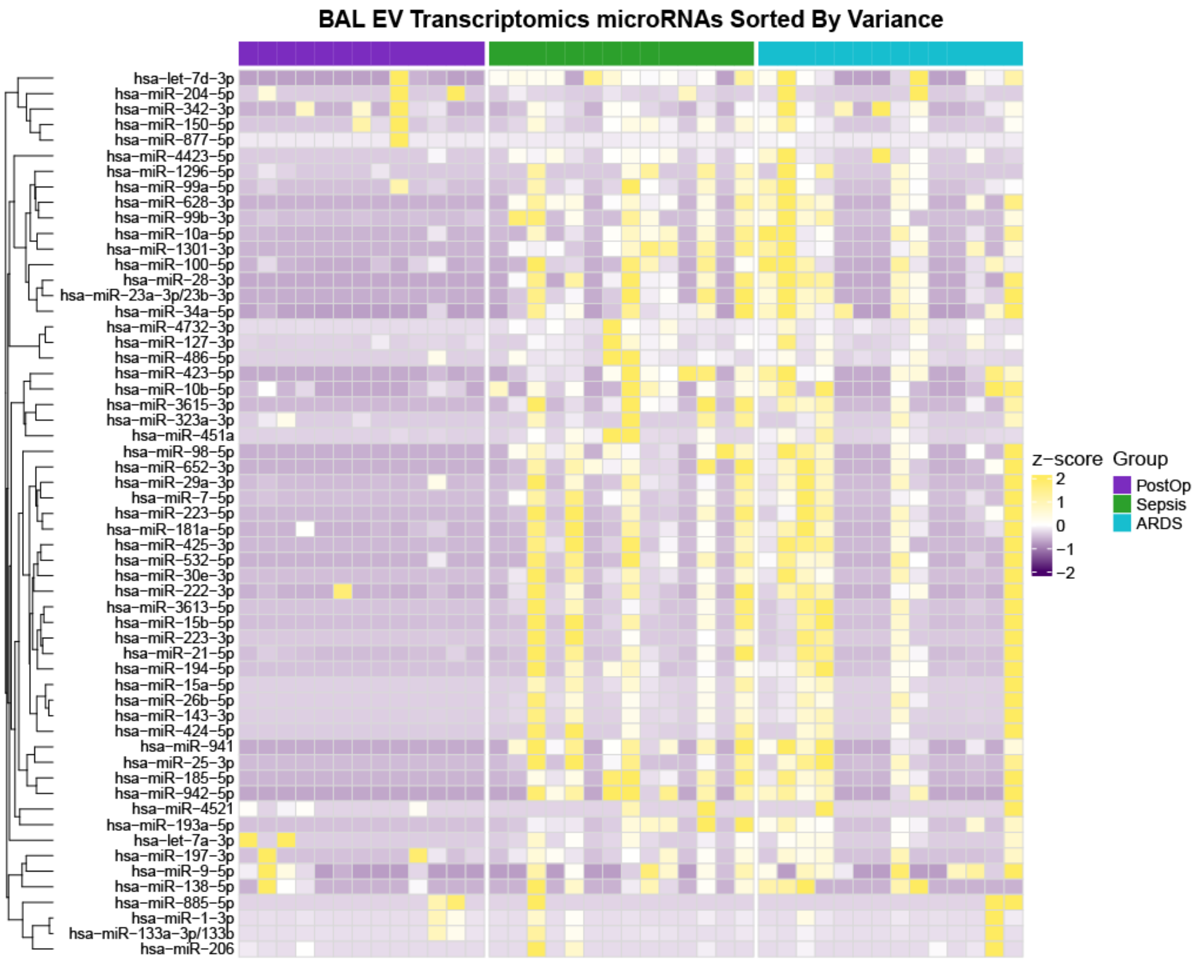


**Supplemental Figure 5A:** Heatmap of differentially expressed BAL EV microRNAs across sepsis patients with and without ARDS, and post-operative controls.

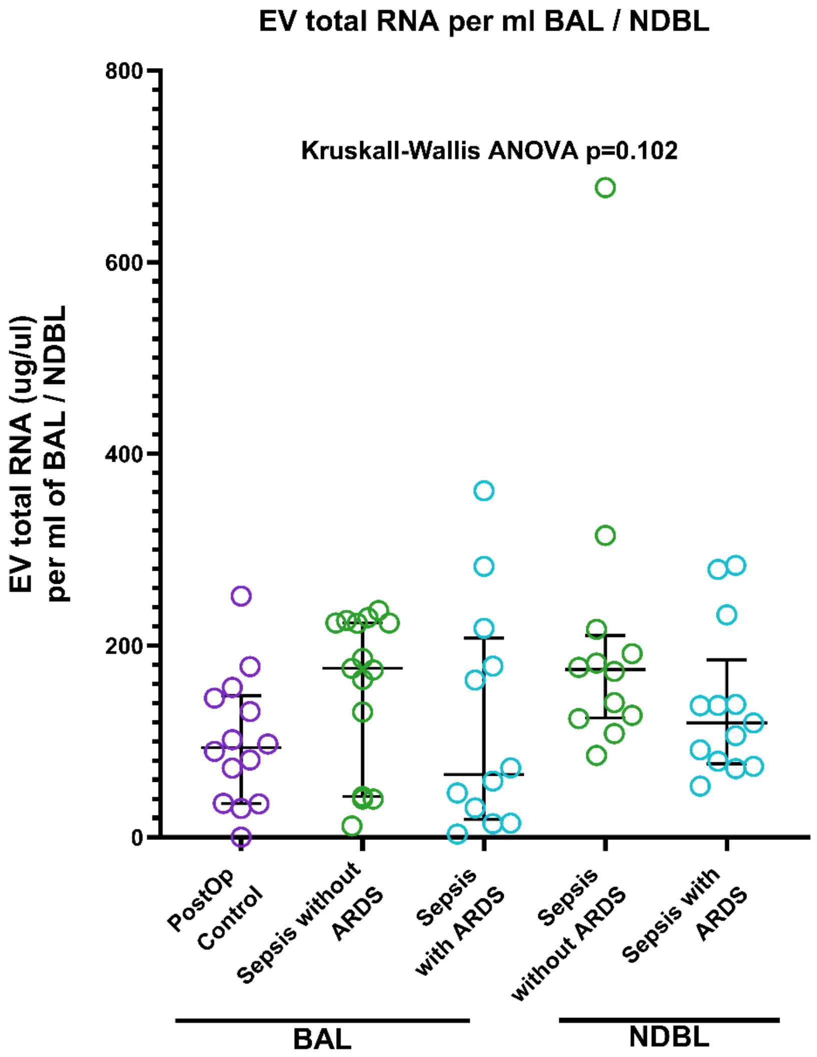


**Supplemental Figure 5B:** Total EV RNA per ml of BAL / NDBL fluid across all patient groups, n≥12.

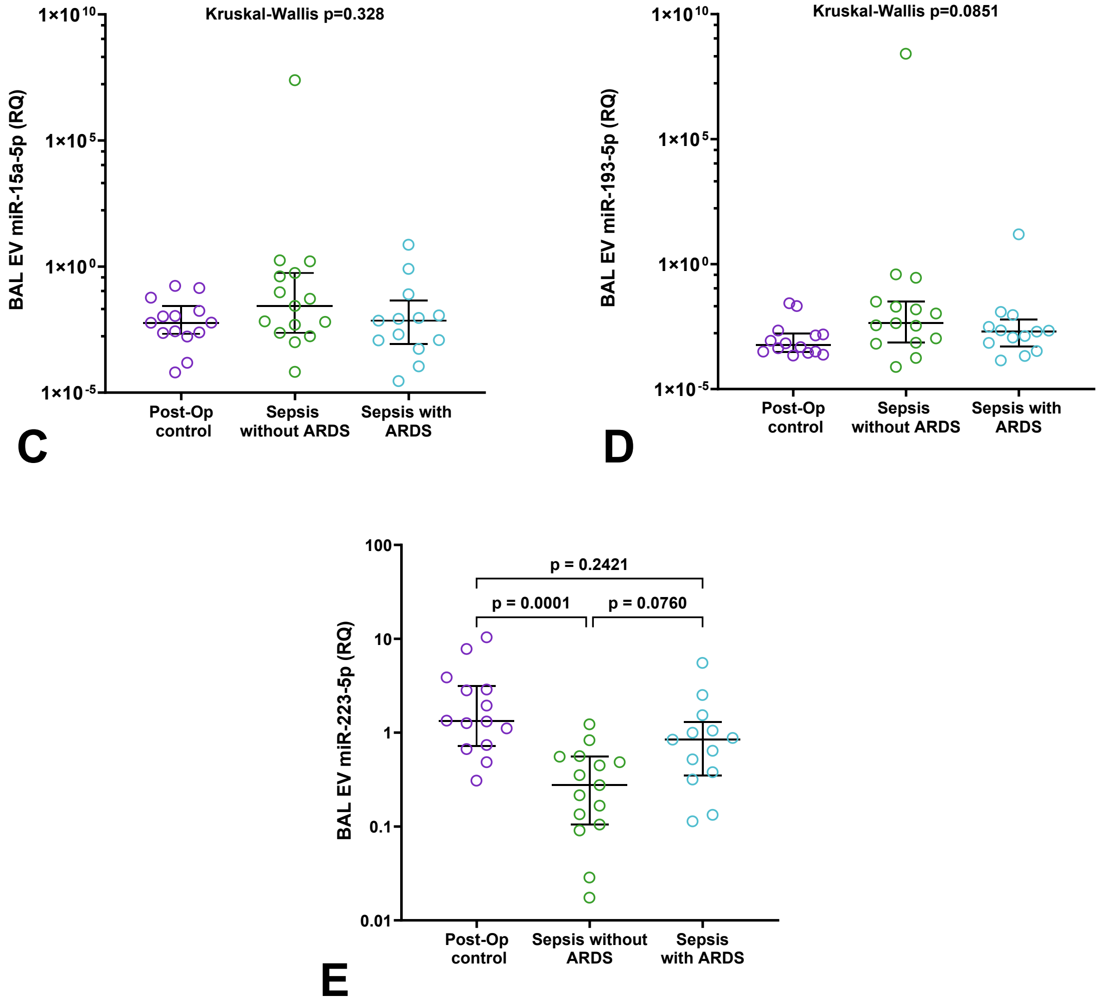


**Supplemental Figure 5C-E:** RT-qPCR of miR-15a-5p, miR193-5p and miR-223-5p from BAL EVs of sepsis patients with and without ARDS and postoperative controls, n≥13. BAL: Broncho-alveolar lavage, EV: Extracellular vesicle.

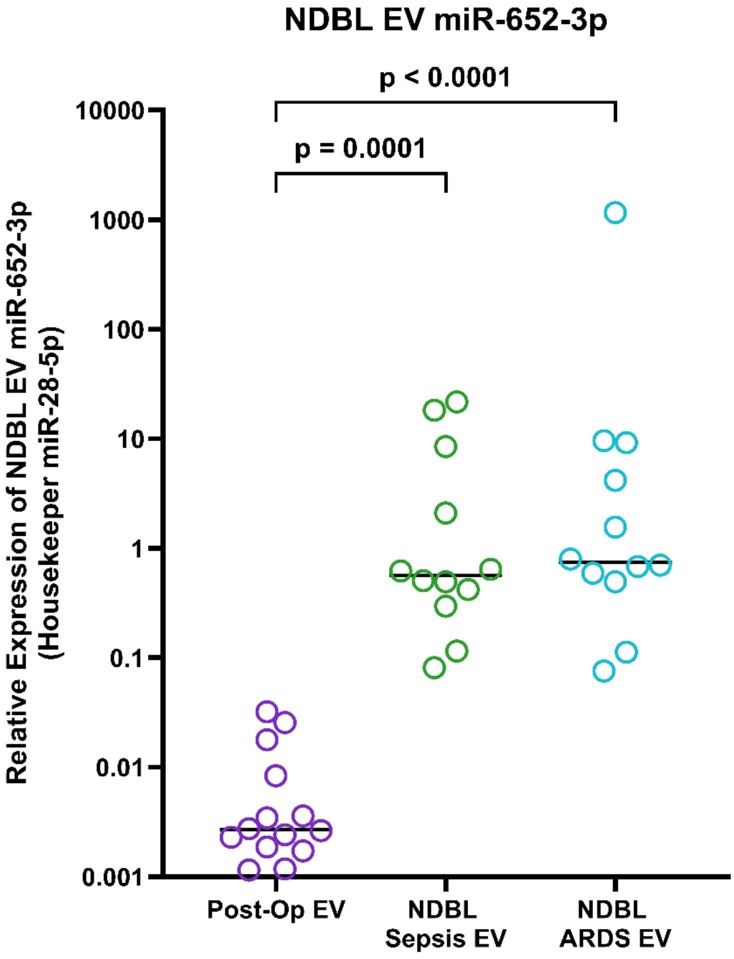


**Supplemental Figure 5F:** RT-qPCR of miR-652-3p in the NDBL EVs of EVs of sepsis patients with and without ARDS and BAL EVs of postoperative controls, n≥12. EV: Extracellular vesicle, NDBL: Non-directed bronchial lavage.

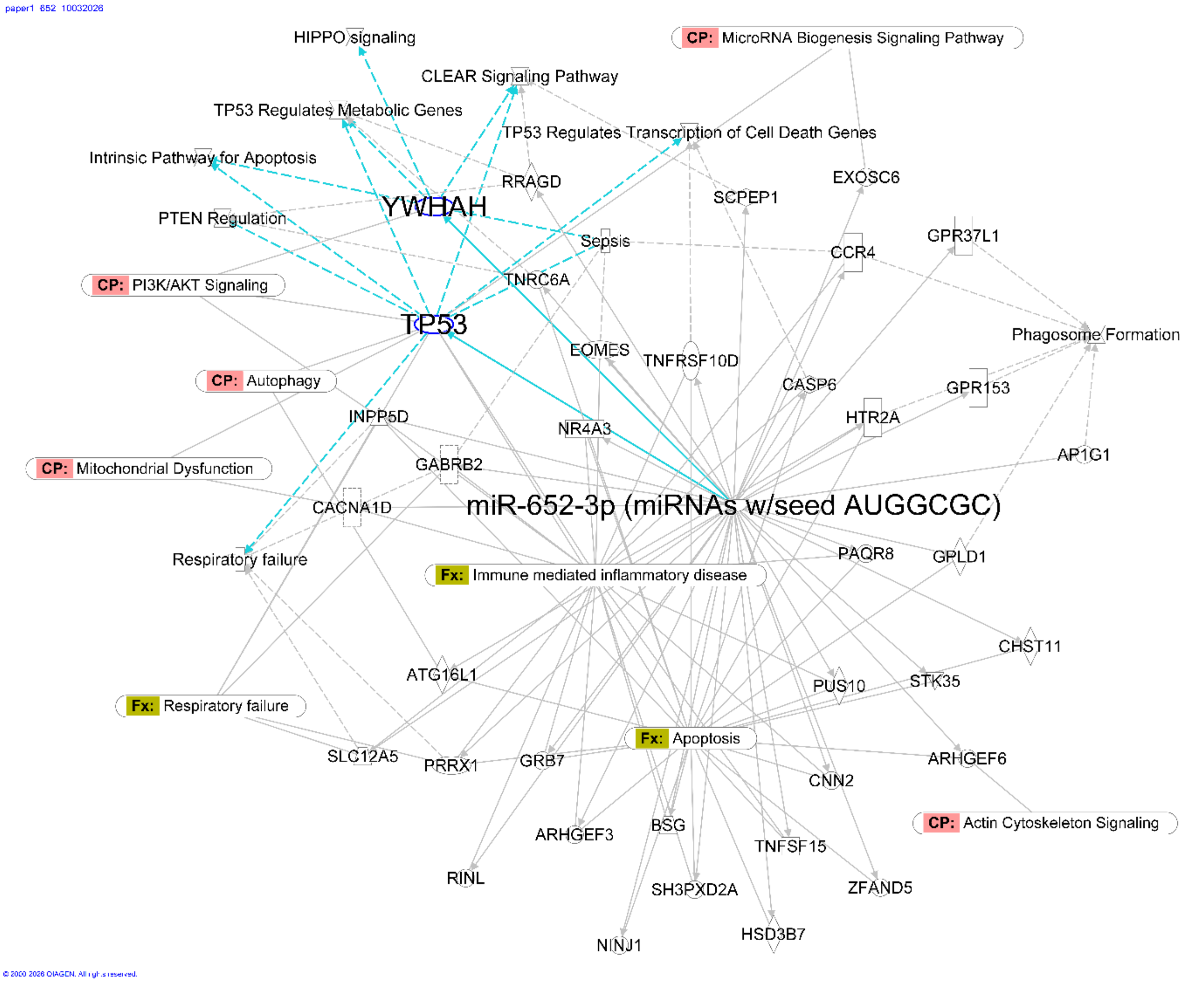


**Supplemental Figure 5G:** Ingenuity Pathway Analysis (Qiagen) target filter network diagram for miR-652-3p

### SUPPLEMENTARY TABLES

**Supplementary Table 1:** Small RNA abundance and read information from BAL EV small RNA-Seq

| Sample name | Trimmed Total Reads | miRNA Reads (%) | mature tRNA Reads (%) | snoRNA Reads (%) | rRNA Reads (%) | ncRNA others (%) | mRNA Reads (%) | Remaining Reads (%) |
|---|---|---|---|---|---|---|---|---|
| Postop_1 | 23667243 | 4058493 (17.15) | 34112 (0.14) | 8896 (0.04) | 345263 (1.46) | 351159 (1.48) | 11525 (0.05) | 18857707 (79.68) |
| Postop_2 | 25118297 | 713276 (2.84) | 285047 (1.13) | 34524 (0.14) | 689971 (2.75) | 1418830 (5.65) | 206827 (0.82) | 21765197 (86.65) |
| Postop_3 | 25246067 | 260673 (1.03) | 145432 (0.58) | 571 (0.00) | 1705689 (6.76) | 92565 (0.37) | 627 (0.00) | 23040397 (91.26) |
| Postop_4 | 24831954 | 979849 | 65310 | 7310 | 1373792 | 173844 | 10394 | 22221383 |

| | | | | | | | | |
|---|---|---|---|---|---|---|---|---|
| | | (3.95) | (0.26) | (0.03) | (5.53) | (0.70) | (0.04) | (89.49) |
| Postop_5 | 24994327 | 392972 | 69036 | 36226 | 2216217 | 313225 | 8454 | 21958191 |
| | | (1.57) | (0.28) | (0.14) | (8.87) | (1.25) | (0.03) | (87.85) |
| Postop_6 | 24960703 | 645849 | 104837 | 3833 | 1815613 | 107989 | 1633 | 22280940 |
| | | (2.59) | (0.42) | (0.02) | (7.27) | (0.43) | (0.01) | (89.26) |
| Postop_7 | 25121401 | 769525 | 1223551 | 6290 | 5490224 | 465111 | 91326 | 17075366 |
| | | (3.06) | (4.87) | (0.03) | (21.85) | (1.85) | (0.36) | (67.97) |
| Postop_8 | 21984203 | 2860625 | 7716 | 10657 | 67487 | 1395288 | 44006 | 17597861 |
| | | (13.01) | (0.04) | (0.05) | (0.31) | (6.35) | (0.20) | (80.05) |
| Postop_9 | 25023102 | 2781804 | 1869561 | 24544 | 3954754 | 880914 | 170504 | 15322753 |
| | | (11.12) | (7.47) | (0.10) | (15.80) | (3.52) | (0.68) | (61.23) |
| Postop_10 | 25220782 | 585476 | 243802 | 72487 | 645059 | 816399 | 52324 | 22801356 |
| | | (2.32) | (0.97) | (0.29) | (2.56) | (3.24) | (0.21) | (90.41) |
| Postop_11 | 25330125 | 268198 | 2264202 | 13397 | 1571485 | 1602515 | 240041 | 19364974 |
| | | (1.06) | (8.94) | (0.05) | (6.20) | (6.33) | (0.95) | (76.45) |

| | | | | | | | | |
|---|---|---|---|---|---|---|---|---|
| Postop_12 | 24942810 | 530791 | 528911 | 42672 | 2899594 | 365970 | 10676 | 20564192 |
| | | (2.13) | (2.12) | (0.17 | 11.62 | 1.47 | 0.04 | 82.45 |
| Postop_13 | 25134737 | 177414 | 6777 | 8903 | 20232 | 135917 | 5751 | 24779455 |
| | | (0.71) | (0.03) | (0.04) | (0.08) | (0.54) | (0.02) | (98.59) |
| ARDS_1 | 24815227 | 3868657 | 286858 | 61176 | 2402003 | 2272408 | 224850 | 15699253 |
| | | (15.59) | (1.16) | (0.25) | (9.68) | (9.16) | (0.91) | (63.26) |
| ARDS_2 | 23266066 | 1049367 | 1109147 | 39270 | 937727 | 3439824 | 458361 | 16225867 |
| | | (4.51) | (4.77) | (0.17) | (4.03) | (14.78) | (1.97) | (69.74) |
| ARDS_3 | 27510997 | 4462179 | 2617875 | 62254 | 1500874 | 4602006 | 460937 | 13754786 |
| | | (16.22) | (9.52) | (0.23) | (5.46) | (16.73) | (1.68) | (50.00) |
| ARDS_4 | 27525951 | 1342000 | 1110960 | 59486 | 517479 | 3539562 | 515974 | 20422218 |
| | | (4.88) | (4.04) | (0.22) | (1.88) | (12.86) | (1.87) | (74.19) |
| ARDS_5 | 27842816 | 2407197 | 4578766 | 29606 | 2586778 | 5258463 | 437403 | 12523405 |
| | | (8.65) | (16.45) | (0.11) | (9.29) | (18.89) | (1.57) | (44.98) |
| ARDS_6 | 27059638 | 1404274 | 1136630 | 14726 | 1300574 | 1755737 | 178765 | 21245839 |

| | | | | | | | | |
|---|---|---|---|---|---|---|---|---|
| | | (5.19) | (4.20) | (0.05) | (4.81) | (6.49) | (0.66) | (78.51) |
| ARDS_7 | 27771222 | 2822166 | 2650659 | 20330 | 501070 | 6480628 | 329341 | 14954913 |
| | | (10.16) | (9.54) | (0.07) | (1.80) | (23.34) | (1.19) | (53.85) |
| ARDS_8 | 27787079 | 5613783 | 4661919 | 17083 | 813155 | 11004473 | 167768 | 5479932 |
| | | (20.20) | (16.78) | (0.06) | (2.93) | (39.60) | (0.60) | (19.72) |
| ARDS_9 | 22810798 | 1537109 | 309966 | 56253 | 1423678 | 4030775 | 328162 | 15112724 |
| | | (6.74) | (1.36) | (0.25) | (6.24) | (17.67) | (1.44) | (66.25) |
| ARDS_10 | 21635342 | 1482304 | 148051 | 51140 | 207525 | 4495029 | 431303 | 14798449 |
| | | (6.85) | (0.68) | (0.24) | (0.96) | (20.78) | (1.99) | (68.40) |
| ARDS_11 | 26735203 | 876991 | 229641 | 133172 | 790442 | 3274289 | 377451 | 20985568 |
| | | (3.28) | (0.86) | (0.50) | (2.96) | (12.25) | (1.41) | (78.49) |
| ARDS_12 | 27631603 | 2472743 | 2313137 | 30010 | 712226 | 10579164 | 293440 | 11215778 |
| | | (8.95) | (8.37) | (0.11) | (2.58) | (38.29) | (1.06) | (40.59) |
| ARDS_13 | 27165286 | 1888107 | 883500 | 30864 | 2130714 | 873739 | 97412 | 21243299 |
| | | (6.95) | (3.25) | (0.11) | (7.84) | (3.22) | (0.36) | (78.20) |

| | | | | | | | | |
|---|---|---|---|---|---|---|---|---|
| ARDS_14 | 27537700 | 3562776 (12.94) | 1337860 (4.86) | 47067 (0.17) | 869716 (3.16) | 10714559 (38.91) | 323812 (1.18) | 10668825 (38.74) |
| Sepsis_1 | 24066883 | 2009117 (8.35) | 775831 (3.22) | 15320 (0.06) | 248003 (1.03) | 2344017 (9.74) | 462150 (1.92) | 18207051 (75.65) |
| Sepsis_2 | 27574205 | 6381550 (23.14) | 3146214 (11.41) | 30600 (0.11) | 1423606 (5.16) | 6217670 (22.55) | 250076 (0.91) | 10107050 (36.65) |
| Sepsis_3 | 27760968 | 2666031 (9.60) | 2151013 (7.75) | 46791 (0.17) | 1687715 (6.08) | 10269636 (36.99) | 315018 (1.13) | 10596824 (38.17) |
| Sepsis_4 | 27744240 | 3046201 (10.98) | 1822840 (6.57) | 32849 (0.12) | 2235202 (8.06) | 4168826 (15.03) | 331514 (1.19) | 16092549 (58.00) |
| Sepsis_5 | 23157261 | 1825919 (7.88) | 125592 (0.54) | 353 (0.00) | 1196777 (5.17) | 453357 (1.96) | 51598 (0.22) | 19503597 (84.22) |
| Sepsis_6 | 27028443 | 2276927 (8.42) | 49627 (0.18) | 2856 (0.01) | 305726 (1.13) | 461251 (1.71) | 16084 (0.06) | 23915847 (88.48) |
| Sepsis_7 | 27560212 | 434212 | 118843 | 51972 | 1441742 | 1818685 | 222682 | 23451692 |

| | | | | | | | | |
|---|---|---|---|---|---|---|---|---|
| | | (1.58) | (0.43) | (0.19) | (5.23) | (6.60) | (0.81) | (85.09) |
| Sepsis_8 | 27698316 | 3277018 | 875256 | 23250 | 371243 | 3138753 | 206601 | 19798270 |
| | | (11.83) | (3.16) | (0.08) | (1.34) | (11.33) | (0.75) | (71.48) |
| Sepsis_9 | 27475117 | 2138381 | 3391868 | 43346 | 856750 | 2352167 | 509779 | 18174432 |
| | | (7.78) | (12.35) | (0.16) | (3.12) | (8.56) | (1.86) | (66.15) |
| Sepsis_10 | 27304382 | 5069592 | 5567 | 1491 | 66599 | 184844 | 7072 | 21969135 |
| | | (18.57) | (0.02) | (0.01) | (0.24) | (0.68) | (0.03) | (80.46) |
| Sepsis_11 | 23142933 | 3440776 | 67652 | 314 | 1022000 | 1162635 | 70586 | 17378927 |
| | | (14.87) | (0.29) | (0.00) | (4.42) | (5.02) | (0.31) | (75.09) |
| Sepsis_12 | 26916266 | 1355451 | 113944 | 109107 | 163383 | 3120584 | 561617 | 21460491 |
| | | (5.04) | (0.42) | (0.41) | (0.61) | (11.59) | (2.09) | (79.73) |
| Sepsis_13 | 27756867 | 2249560 | 117006 | 55722 | 2164930 | 3063353 | 432658 | 19659972 |
| | | (8.10) | (0.42) | (0.20) | (7.80) | (11.04) | (1.56) | (70.83) |
| Sepsis_14 | 25656410 | 5056723 | 1983320 | 38620 | 1685294 | 8867956 | 235890 | 7766864 |
| | | (19.71) | (7.73) | (0.15) | (6.57) | (34.56) | (0.92) | (30.27) |

**Supplementary Table 2:** List of differentially expressed BAL EV miRNAs among post-operative controls (n=13) and sepsis patients with ARDS (n=14) assessed by small RNA-Seq, ranked by p value (<0.05)

| miRNA | Mean Postop Control (reads) | Mean ARDS (reads) | Absolute Change | Log2 Fold Change | p-value | FDR (p-adjusted) |
|---|---|---|---|---|---|---|
| hsa-miR-652-3p | 1.076923077 | 9436.285714 | 9435.208791 | 6.248 | 8.10E-13 | 2.42E-10 |
| hsa-miR-15a-5p | 0.384615385 | 3023.714286 | 3023.32967 | 5.131 | 9.89E-08 | 9.98E-06 |
| hsa-miR-193a-5p | 0 | 3040.5 | 3040.5 | 5.954 | 1.06E-07 | 9.98E-06 |
| hsa-miR-223-5p | 0 | 3603.571429 | 3603.571429 | 5.369 | 1.34E-07 | 9.98E-06 |
| hsa-miR-100-5p | 646.3076923 | 4173.142857 | 3526.835165 | -3.652 | 6.36E-07 | 3.80E-05 |
| hsa-miR-342-3p | 4244.923077 | 2171.571429 | -2073.351648 | -4.378 | 2.36E-06 | 0.00010069 |
| hsa-miR-941 | 0.153846154 | 1523.642857 | 1523.489011 | 4.964 | 2.05E-06 | 0.00010069 |

| | | | | | | |
|---|---|---|---|---|---|---|
| hsa-miR-185-5p | 0.538461538 | 1118.785714 | 1118.247253 | 4.639 | 3.51E-06 | 0.00013126 |
| hsa-miR-302b-3p | 6.538461538 | 0 | -6.538461538 | -13.182 | 6.48E-06 | 0.00021541 |
| hsa-miR-302a-3p | 3.461538462 | 0 | -3.461538462 | -12.205 | 1.66E-05 | 0.00044978 |
| hsa-miR-302d-3p | 3.307692308 | 0 | -3.307692308 | -12.213 | 1.81E-05 | 0.00044978 |
| hsa-miR-3615-3p | 0 | 1170.142857 | 1170.142857 | 4.416 | 1.75E-05 | 0.00044978 |
| hsa-miR-204-5p | 2362.307692 | 563.1428571 | -1799.164835 | -3.770 | 2.47E-05 | 0.00052799 |
| hsa-miR-206 | 6.307692308 | 69.21428571 | 62.90659341 | -7.539 | 2.42E-05 | 0.00052799 |
| hsa-miR-133a-3p/133b | 77.76923077 | 91.35714286 | 13.58791209 | -6.419 | 2.91E-05 | 0.00055578 |
| hsa-miR-181a-5p | 1462.384615 | 12664 | 11201.61538 | -3.403 | 2.97E-05 | 0.00055578 |
| hsa-miR-877-5p | 2132.615385 | 59.5 | -2073.115385 | -6.615 | 3.17E-05 | 0.00055817 |
| hsa-let-7a-3p | 2729.615385 | 990.1428571 | -1739.472527 | -5.681 | 4.10E-05 | 0.00062808 |
| hsa-miR-223-3p | 212.3076923 | 98378.71429 | 98166.40659 | 4.990 | 4.20E-05 | 0.00062808 |
| hsa-miR-4521 | 2 | 7.571428571 | 5.571428571 | -8.128 | 4.10E-05 | 0.00062808 |
| hsa-miR-302c-3p | 4.692307692 | 0 | -4.692307692 | -11.297 | 0.00011353 | 0.00159312 |

| | | | | | | |
|---|---|---|---|---|---|---|
| hsa-miR-98-5p | 1.923076923 | 1649.928571 | 1648.005495 | 3.753 | 0.00011722 | 0.00159312 |
| hsa-miR-30e-3p | 0.384615385 | 1239.714286 | 1239.32967 | 4.015 | 0.00019717 | 0.00245644 |
| hsa-miR-451a | 5121.307692 | 376138 | 371016.6923 | 3.013 | 0.00019193 | 0.00245644 |
| hsa-miR-628-3p | 0 | 562.2857143 | 562.2857143 | 4.003 | 0.00027676 | 0.00330999 |
| hsa-miR-9-5p | 16.76923077 | 24.07142857 | 7.302197802 | -5.018 | 0.00029987 | 0.00344848 |
| hsa-miR-1301-3p | 0 | 408.4285714 | 408.4285714 | 3.866 | 0.00048168 | 0.00533412 |
| hsa-miR-10b-5p | 6.461538462 | 34.57142857 | 28.10989011 | -3.925 | 0.00052351 | 0.00559038 |
| hsa-miR-197-3p | 511.7692308 | 306 | -205.7692308 | -3.947 | 0.00072717 | 0.00749733 |
| hsa-miR-138-5p | 25.76923077 | 43.57142857 | 17.8021978 | -4.556 | 0.00126851 | 0.01264283 |
| hsa-miR-127-3p | 25.46153846 | 386.5714286 | 361.1098901 | -3.524 | 0.00133598 | 0.0128857 |
| hsa-miR-34a-5p | 280.6153846 | 4289.857143 | 4009.241758 | -2.138 | 0.0015655 | 0.01462766 |
| hsa-miR-25-3p | 8.307692308 | 8620 | 8611.692308 | 2.547 | 0.00182722 | 0.0165557 |
| hsa-miR-1-3p | 68.38461538 | 53.78571429 | -14.5989011 | -4.950 | 0.00205686 | 0.01808828 |
| hsa-miR-150-5p | 3038.153846 | 1921.714286 | -1116.43956 | -2.954 | 0.00240148 | 0.02051549 |

| | | | | | | |
|---|---|---|---|---|---|---|
| hsa-miR-23a-3p/23b-3p | 1104.153846 | 139338.6429 | 138234.489 | 1.776 | 0.00265468 | 0.02204859 |
| hsa-miR-29a-3p | 188.8461538 | 1597.357143 | 1408.510989 | -2.480 | 0.00273167 | 0.02207488 |
| hsa-miR-3613-5p | 0.769230769 | 2700.857143 | 2700.087912 | 4.064 | 0.00295961 | 0.02297927 |
| hsa-miR-425-3p | 3.461538462 | 1576.071429 | 1572.60989 | 3.765 | 0.00303072 | 0.02297927 |
| hsa-miR-4732-3p | 0.384615385 | 302.4285714 | 302.043956 | 3.565 | 0.003151 | 0.02297927 |
| hsa-miR-942-5p | 0 | 730.3571429 | 730.3571429 | 4.089 | 0.00310924 | 0.02297927 |
| hsa-miR-935 | 0.923076923 | 6.071428571 | 5.148351648 | -5.549 | 0.00343001 | 0.02441841 |
| hsa-miR-222-3p | 1420 | 5491.928571 | 4071.928571 | -2.109 | 0.00359981 | 0.02503124 |
| hsa-miR-323a-3p | 1.076923077 | 10.42857143 | 9.351648352 | -4.745 | 0.00409662 | 0.02783837 |
| hsa-let-7d-3p | 1080.923077 | 4429.071429 | 3348.148352 | 3.202 | 0.00449254 | 0.02985042 |
| hsa-miR-7-5p | 51.15384615 | 3551.071429 | 3499.917582 | -2.715 | 0.00524782 | 0.03338505 |
| hsa-miR-885-5p | 1.384615385 | 1.785714286 | 0.401098901 | -8.186 | 0.00523721 | 0.03338505 |
| hsa-miR-99b-3p | 4.846153846 | 182.8571429 | 178.010989 | -3.948 | 0.00586131 | 0.03651108 |
| hsa-miR-1296-5p | 1.461538462 | 19.14285714 | 17.68131868 | -4.846 | 0.00599199 | 0.03656338 |

| | | | | | | |
|---|---|---|---|---|---|---|
| hsa-miR-21-5p | 7318.538462 | 150908.4286 | 143589.8901 | -1.916 | 0.0072119 | 0.04228151 |
| hsa-miR-99a-5p | 6219.384615 | 23250.64286 | 17031.25824 | -1.602 | 0.00710262 | 0.04228151 |
| hsa-miR-194-5p | 0.153846154 | 386.7857143 | 386.6318681 | 3.263 | 0.00752046 | 0.04324265 |
| hsa-miR-532-5p | 54.69230769 | 1346.642857 | 1291.950549 | 2.501 | 0.00814787 | 0.04596627 |
| hsa-miR-125b-1-3p | 0.923076923 | 14.07142857 | 13.14835165 | -4.966 | 0.00840998 | 0.04656638 |
| hsa-miR-28-3p | 2.076923077 | 1988.642857 | 1986.565934 | 3.297 | 0.00903625 | 0.04878674 |
| hsa-miR-424-5p | 10.84615385 | 5629.5 | 5618.653846 | 3.211 | 0.00930048 | 0.04878674 |
| hsa-miR-767-3p | 0.461538462 | 0 | -0.461538462 | -6.647 | 0.00920635 | 0.04878674 |
| hsa-miR-190b-5p | 743.3076923 | 2766.642857 | 2023.335165 | 3.278 | 0.01120522 | 0.05776483 |
| hsa-miR-30a-3p | 0.846153846 | 815.1428571 | 814.2967033 | 2.617 | 0.01255297 | 0.06361588 |
| hsa-miR-30d-5p | 538.0769231 | 2835.285714 | 2297.208791 | -1.992 | 0.01533668 | 0.07642777 |
| hsa-miR-511-5p | 1251.307692 | 33.78571429 | -1217.521978 | -3.953 | 0.0160203 | 0.07774635 |
| hsa-miR-92b-3p | 121212.2308 | 255016.7857 | 133804.5549 | -2.474 | 0.01612132 | 0.07774635 |
| hsa-miR-654-3p | 0.384615385 | 2.714285714 | 2.32967033 | -6.130 | 0.01772158 | 0.08410719 |
| hsa-miR-381-3p | 0.230769231 | 2.785714286 | 2.554945055 | -5.743 | 0.01895907 | 0.08857443 |

| | | | | | | |
|---|---|---|---|---|---|---|
| hsa-miR-29c-5p | 15.38461538 | 922.6428571 | 907.2582418 | 2.774 | 0.02042052 | 0.09393441 |
| hsa-let-7i-3p | 1 | 40 | 39 | -4.313 | 0.02074385 | 0.09397594 |
| hsa-miR-103a-3p/107 | 0.538461538 | 1363 | 1362.461538 | 2.943 | 0.02129851 | 0.09504855 |
| hsa-miR-181b-5p | 380.2307692 | 3859.285714 | 3479.054945 | -1.936 | 0.02221154 | 0.09766545 |
| hsa-miR-625-5p | 12.23076923 | 997.3571429 | 985.1263736 | 2.607 | 0.0270879 | 0.1173809 |
| hsa-let-7f-5p | 563.9230769 | 16984.07143 | 16420.14835 | 1.588 | 0.03818764 | 0.1585848 |
| hsa-miR-449a/449b-5p | 3641.615385 | 20267.5 | 16625.88462 | -2.135 | 0.03795501 | 0.1585848 |
| hsa-miR-582-5p | 0 | 677.7857143 | 677.7857143 | 2.932 | 0.03808089 | 0.1585848 |
| hsa-miR-126-3p | 37.61538462 | 1406.928571 | 1369.313187 | -1.922 | 0.03951167 | 0.1614056 |
| hsa-miR-214-3p | 1.153846154 | 16 | 14.84615385 | -3.443 | 0.03994654 | 0.1614056 |
| hsa-miR-205-5p | 77.76923077 | 1001.285714 | 923.5164835 | 3.241 | 0.04184786 | 0.16683349 |
| hsa-let-7e-3p | 2.461538462 | 133.2142857 | 130.7527473 | -2.842 | 0.04305557 | 0.16938969 |
| hsa-miR-423-5p | 22.30769231 | 1348.285714 | 1325.978022 | 2.265 | 0.04379209 | 0.17004981 |
| hsa-let-7a-5p/7c-5p | 8032.230769 | 66453.85714 | 58421.62637 | 1.647 | 0.04494322 | 0.17228233 |

| | | | | | | |
|---|---|---|---|---|---|---|
| hsa-miR-26b-5p | 21.76923077 | 12239.14286 | 12217.37363 | 1.871 | 0.04635135 | 0.17349375 |
| hsa-miR-379-5p | 0.153846154 | 12 | 11.84615385 | -3.747 | 0.04641973 | 0.17349375 |
| hsa-miR-10401-3p | 5.307692308 | 18.07142857 | 12.76373626 | -4.242 | 0.04764496 | 0.1758746 |

**Supplementary Table 3:** List of differentially expressed BAL EV miRNAs among post-operative controls (n=13) and sepsis patients without ARDS (n=14) assessed by small RNA-Seq, ranked by p value (<0.05)

| miRNA | Mean Control (reads) | Mean Sepsis | Absolute Change | Log2 Fold Change | p-value | p-adjusted (FDR) |
|---|---|---|---|---|---|---|
| hsa-miR-652-3p | 1.076923077 | 8630.5 | 8629.423077 | 6.382 | 2.20E-13 | 3.96E-11 |
| hsa-miR-223-5p | 0 | 3316.428571 | 3316.428571 | 5.767 | 2.70E-08 | 2.43E-06 |
| hsa-let-7a-3p | 2729.615385 | 1077.642857 | -1651.972527 | -9.251 | 5.79E-08 | 3.4742E-06 |
| hsa-miR-15a-5p | 0.384615385 | 4609.285714 | 4608.901099 | 5.967 | 5.10E-07 | 2.30E-05 |

| | | | | | | |
|---|---|---|---|---|---|---|
| hsa-miR-877-5p | 2132.615385 | 78.42857143 | -2054.186813 | -7.524 | 2.76E-06 | 9.9469E-05 |
| hsa-miR-204-5p | 2362.307692 | 1414.642857 | -947.6648352 | -4.567 | 4.14E-06 | 0.00012427 |
| hsa-miR-342-3p | 4244.923077 | 5440.714286 | 1195.791209 | -4.338 | 7.76E-06 | 0.00019943 |
| hsa-miR-21-5p | 7318.538462 | 122461.0714 | 115142.533 | -4.000 | 1.3694E-05 | 0.00030525 |
| hsa-miR-193a-5p | 0 | 1531.785714 | 1531.785714 | 5.419 | 1.53E-05 | 3.05E-04 |
| hsa-miR-23a-3p/23b-3p | 1104.153846 | 141070.4286 | 139966.2747 | 2.126 | 1.70049E-05 | 0.00030609 |
| hsa-miR-223-3p | 212.3076923 | 107883.2143 | 107670.9066 | 5.649 | 2.76E-05 | 0.00045139 |
| hsa-miR-10b-5p | 6.461538462 | 34.28571429 | 27.82417582 | -4.134 | 0.000102288 | 0.00153432 |
| hsa-miR-143-3p | 33.61538462 | 18224.42857 | 18190.81319 | 3.397 | 0.000421145 | 0.00583124 |
| hsa-miR-3615-3p | 0 | 636.8571429 | 636.8571429 | 4.111 | 5.16E-04 | 0.00618371 |
| hsa-miR-194-5p | 0.153846154 | 542.2142857 | 542.0604396 | 5.160 | 0.000531005 | 0.00618371 |
| hsa-miR-302b-3p | 6.538461538 | 0 | -6.538461538 | -9.862 | 5.50E-04 | 0.00618371 |
| hsa-miR-185-5p | 0.538461538 | 886.2142857 | 885.6758242 | 3.882 | 8.49E-04 | 0.00820503 |
| hsa-miR-3613-5p | 0.769230769 | 3705.214286 | 3704.445055 | 4.673 | 0.000852036 | 0.00820503 |

| | | | | | | |
|---|---|---|---|---|---|---|
| hsa-miR-150-5p | 3038.153846 | 1594.428571 | -1443.725275 | -3.007 | 0.000866086 | 0.00820503 |
| hsa-miR-133a-3p/133b | 77.76923077 | 279.4285714 | 201.6593407 | -5.475 | 9.55E-04 | 0.00859188 |
| hsa-miR-302d-3p | 3.307692308 | 0 | -3.307692308 | -9.047 | 1.21E-03 | 0.00991941 |
| hsa-miR-302a-3p | 3.461538462 | 0 | -3.461538462 | -8.940 | 1.21E-03 | 0.00991941 |
| hsa-miR-222-3p | 1420 | 5489.428571 | 4069.428571 | -2.223 | 0.001413545 | 0.01106252 |
| hsa-miR-942-5p | 0 | 446.7857143 | 446.7857143 | 4.139 | 0.001560985 | 0.01170739 |
| hsa-miR-4423-5p | 72 | 2305.285714 | 2233.285714 | 4.661 | 0.001640405 | 0.01181091 |
| hsa-miR-30e-3p | 0.384615385 | 1302.5 | 1302.115385 | 3.784 | 0.001743057 | 0.01206731 |
| hsa-miR-941 | 0.153846154 | 1106.071429 | 1105.917582 | 3.903 | 2.13E-03 | 0.01420366 |
| hsa-miR-15b-5p | 38.69230769 | 16446 | 16407.30769 | 2.302 | 0.002706106 | 0.0173964 |
| hsa-miR-197-3p | 511.7692308 | 303.5 | -208.2692308 | -3.831 | 0.00282454 | 0.01753162 |
| hsa-miR-26b-5p | 21.76923077 | 22200.57143 | 22178.8022 | 2.993 | 0.003510197 | 0.02106118 |
| hsa-miR-302c-3p | 4.692307692 | 0 | -4.692307692 | -8.363 | 0.004081941 | 0.02370159 |
| hsa-miR-486-5p | 111 | 534.7857143 | 423.7857143 | 3.712 | 0.004577085 | 0.0257461 |

| | | | | | | |
|---|---|---|---|---|---|---|
| hsa-miR-99b-3p | 4.846153846 | 186.2142857 | 181.3681319 | -4.415 | 0.00485308 | 0.02647135 |
| hsa-miR-425-3p | 3.461538462 | 1609.5 | 1606.038462 | 4.166 | 0.00576826 | 0.03053785 |
| hsa-miR-181a-5p | 1462.384615 | 15294 | 13831.61538 | -1.580 | 6.34E-03 | 0.0325998 |
| hsa-miR-10a-5p | 25 | 625.1428571 | 600.1428571 | 2.744 | 0.006733534 | 0.03336374 |
| hsa-miR-323a-3p | 1.076923077 | 3.785714286 | 2.708791209 | -4.597 | 0.006858102 | 0.03336374 |
| hsa-miR-628-3p | 0 | 781.5714286 | 781.5714286 | 3.738 | 0.007796046 | 0.03692864 |
| hsa-miR-138-5p | 25.76923077 | 55.28571429 | 29.51648352 | -4.624 | 0.009868075 | 0.04480209 |
| hsa-miR-423-5p | 22.30769231 | 1251.071429 | 1228.763736 | 2.760 | 0.009956019 | 0.04480209 |
| hsa-miR-25-3p | 8.307692308 | 7112.5 | 7104.192308 | 2.229 | 0.012890927 | 0.05659431 |
| hsa-miR-4521 | 2 | 6.928571429 | 4.928571429 | -5.005 | 1.51E-02 | 0.06181037 |
| hsa-miR-28-3p | 2.076923077 | 2263.214286 | 2261.137363 | 3.164 | 0.015470623 | 0.06181037 |
| hsa-miR-146b-5p | 0.230769231 | 796.0714286 | 795.8406593 | 3.344 | 0.01561359 | 0.06181037 |
| hsa-miR-574-3p | 31.53846154 | 456.7857143 | 425.2472527 | -1.898 | 0.015858044 | 0.06181037 |
| hsa-miR-27a-3p/27b-3p | 1012.538462 | 59557.64286 | 58545.1044 | 2.027 | 0.015897598 | 0.06181037 |

| | | | | | | |
|---|---|---|---|---|---|---|
| hsa-let-7b-5p | 28948.53846 | 124397.5 | 95448.96154 | -2.012 | 0.016139375 | 0.06181037 |
| hsa-miR-7-5p | 51.15384615 | 4900.785714 | 4849.631868 | -2.309 | 0.02098321 | 0.07672156 |
| hsa-miR-98-5p | 1.923076923 | 1634.857143 | 1632.934066 | 2.758 | 0.02125607 | 0.07672156 |
| hsa-miR-369-5p | 2.769230769 | 143.0714286 | 140.3021978 | -2.522 | 0.021311545 | 0.07672156 |
| hsa-let-7f-5p | 563.9230769 | 29155.21429 | 28591.29121 | 1.220 | 0.022482984 | 0.07935171 |
| hsa-miR-29a-3p | 188.8461538 | 2009.071429 | 1820.225275 | -1.981 | 0.023298848 | 0.08064986 |
| hsa-miR-99a-5p | 6219.384615 | 30474.5 | 24255.11538 | -1.281 | 0.025811195 | 0.08532801 |
| hsa-miR-190b-5p | 743.3076923 | 3187.785714 | 2444.478022 | 3.299 | 0.026019823 | 0.08532801 |
| hsa-miR-625-5p | 12.23076923 | 1416.142857 | 1403.912088 | 2.630 | 0.026072446 | 0.08532801 |
| hsa-miR-1301-3p | 0 | 410.2142857 | 410.2142857 | 3.231 | 0.027591483 | 0.08868691 |
| hsa-miR-30d-5p | 538.0769231 | 3931.214286 | 3393.137363 | -1.849 | 0.029806858 | 0.09412692 |
| hsa-miR-92a-3p | 18258.84615 | 69233.21429 | 50974.36813 | -1.479 | 0.031718723 | 0.09843742 |
| hsa-miR-100-5p | 646.3076923 | 5074.357143 | 4428.049451 | -1.797 | 3.27E-02 | 9.97E-02 |
| hsa-miR-99b-5p | 2114.384615 | 9764.571429 | 7650.186813 | -1.245 | 0.035983816 | 0.10795145 |
| hsa-miR-30a-3p | 0.846153846 | 1286.214286 | 1285.368132 | 2.554 | 0.036745525 | 0.10842942 |

| | | | | | | |
|---|---|---|---|---|---|---|
| hsa-miR-92b-3p | 121212.2308 | 136818.2857 | 15606.05495 | -2.246 | 0.037651316 | 0.10931027 |
| hsa-miR-16-5p | 83.92307692 | 36277.21429 | 36193.29121 | 1.661 | 0.040399911 | 0.11542832 |
| hsa-miR-4732-3p | 0.384615385 | 92.5 | 92.11538462 | 3.018 | 0.04355809 | 0.12250713 |
| hsa-miR-214-3p | 1.153846154 | 32.14285714 | 30.98901099 | -3.333 | 0.044778317 | NA |
| hsa-miR-582-5p | 0 | 1097.714286 | 1097.714286 | 3.387 | 0.047812258 | 0.13240318 |

**Supplementary Table 4:** Primers used for microRNA and mRNA RT-qPCR

| Name | Catalogue number | Supplier | Sequence |
|---|---|---|---|
| hsa-miR-652-3p Taqman | 478189_mir | Applied Biosciences | 5'-AAUGGCGCCACUAGGGUUGUG-3' |
| hsa-miR-28-5p Taqman | 478000_mir | Applied Biosciences | 5'-AAGGAGCUCACAGUCUAUUGAG-3' |
| hsa-mir-193a-5p Taqman | 477954_mir | Applied Biosciences | 5'-UGGGUCUUUGCGGGCGAGAUGA-3' |
| hsa-mir-223-5p Taqman | 477984_mir | Applied Biosciences | 5'-CGUGUAUUUGACAAGCUGAGUU-3' |
| hsa-mir-15a-5p Taqman | 477858_mir | Applied Biosciences | 5'-UAGCAGCACAUAAUGGUUUGUG-3' |

| | | | |
|---|---|---|---|
| YWHAH HP206927 qSTAR qpcr Primer Pair | NM_003405 | OriGene | 5'-ACGACATGGCCTCCGCTATGAA-3' |
| P53 qPCr qSTAR Primer Pair | hp200518 | OriGene | 5'-CCTCAGCATCTTATCCGAGTGG-3' |
| Eukaryotic 18S rRNA Endogenous Control | Hs99999901_s1 | Applied Biosciences | Proprietary |

**Supplementary Table 5:** Antibodies used across all methods. All anti-human unless otherwise specified. FC: flow cytometry, ICC: Immunocytochemistry

| **Name and conjugation** | **Supplier** | **Catalogue number** | **Dilution** | **Source** |
|---|---|---|---|---|
| CD14-Allophycocyanin (clone 61D3) | Invitrogen | 17-0149-42 | 1:100 Exoview | Mouse IgG1k |
| EpCam-AlexaFluor488 (clone 94C) | Biolegend | 324210 | 1:400 Exoview | Mouse IgG2b,k |
| CD66b-AlexaFluor647 (clone G10F5) | BD Biosciences | 561645 | 1:200 Exoview | Mouse IgM k |
| LC3B | Cell signalling technologies | 83506S | 51 ug/ml<br>1:500 ICC | Mouse IgG2b |
| CD107a LAMP-1 (clone H4A3) | BioRad laboratories | MCA6113Z | 1.0 mg/ml<br>1:500 ICC | Mouse IgG1 |
| CD63 - APC | Thermofisher Scientific | MA5-30187 | 1mg/ml<br>1:100 ICC | Recombinant Monoclonal Rabbit IgG |
| Anti-rabbit Alexa FluorTM 568 | Thermofisher Scientific | A10042 | 2mg/ml<br>1:250 ICC | Donkey/ IgG |
| Anti-mouse AlexaFluor Plus 488 | Invitrogen | A32723 | 2mg/ml<br>1:500 ICC | Goat /IgG |
| Anti-rabbit IgG Alexafluor Plus 594 | Invitrogen | A32740 | 2mg/ml<br>1:250 ICC | Goat/IgG |

| | | | | |
|---|---|---|---|---|
| TOMM20 | Sigma Aldrich | HPA011562 | 0.20 mg/ml<br>1:500 ICC | Rabbit polyclonal |
| CD68 APC | Biolegend U.K | Y1/82A | 0.2 mg/ml<br>1:100 FC | Rat IgG2a |
